# Genomic profiling of young-onset g*BRCA1/2* breast cancer reveals distinct genomic landscapes and therapeutic implications for PARP and CDK4/6 inhibitor selection

**DOI:** 10.1101/2025.09.19.25336150

**Authors:** Mwangala P. Akamandisa, Mingyi Xia, Wilson Cheah, Bradley Wubbenhorst, Kurt D’Andrea, Mengyao Fan, Jake Shilan, Dana Pueschl, Anupma Nayak, Hayley McKenzie, William Tapper, Ellen R. Copson, Ramsey I. Cutress, Susan M. Domchek, Diana M. Eccles, Katherine L. Nathanson

## Abstract

**Purpose:** g*BRCA1/2* pathogenic variant (PV) carriers have elevated young-onset breast cancer risk. Understanding the distinct genomic landscapes of g*BRCA1-* and g*BRCA2*-associated breast cancer, including presence of PARP and CDK4/6 inhibitor (PARPi; CDK4/6i) response-associated alterations, may inform treatment selection for these patients.

**Patients and methods:** We evaluated 136 treatment-naïve primary tumors from POSH study participants diagnosed with breast cancer before age 50 years (92.6% diagnosed ≤40): g*BRCA1* 86(63.2%), g*BRCA2* 50(36.8%). We evaluated somatic mutational and copy number variations (CNV), allele-specific loss of heterozygosity (asLOH), homologous recombination deficiency (HRD), and single-base substitution signatures (SBS) from whole exome sequencing.

**Results:** Both g*BRCA1* (93%) and g*BRCA2* (96%) breast cancers had high rates of asLOH. We found significant differences between g*BRCA1* and g*BRCA2* tumors in average HRD scores (57.4±1.3 vs 43.7±1.5, p<0.0001) and SBS composition: SBS1 (aging-associated) 12.9 vs 7.3, p=0.013; SBS18 (reactive oxygen species [ROS]-associated) 1.4 vs 0, p=0.007; SBS3 (HRD-associated) 27.3 vs 42.6, p=0.002; and SBS26 (mismatch repair-associated) 5.9 vs 9.4, p=0.049. Compared to g*BRCA2* tumors, g*BRCA1* tumors with asLOH were significantly enriched for gains of chr6q, and alterations in Hallmark ROS, DNA repair, and epithelial-mesenchymal transition pathways. In ER-positive, HER2-negative tumors from POSH g*BRCA1/2* carriers compared to noncarriers from the TCGA, we found significant enrichment of *RB1* (OR:6.3, 95%CI:2.8-15.4, p_adj_=0.001), *TP53* (OR:4.6, 95%CI:1.9-12.1, p_adj_=0.017), *FAT1* (OR:3.9, 95%CI:1.84-8.7, p_adj_=0.013), and *MYC* (OR:4.0, 95%CI:1.8-9.1, p_adj_=0.017) SNV/indels/CNVs, which are associated with CDK4/6i resistance.

**Conclusion:** Our data suggest that PARPi may be preferable over CDK4/6i to treat ER-positive, HER2-negative breast cancer in young-onset g*BRCA1/2*-associated breast cancer when both therapies are considered in adjuvant and metastatic settings. Additionally, we identified significant differences between g*BRCA1-* and g*BRCA2*-associated tumors, which may inform therapeutic development.

## Introduction

Approximately 12% of young-onset breast cancer patients (first diagnosis at age ≤40 years) and 30% of very young-onset breast cancer patients (first diagnosis at age ≤30 years) carry germline *BRCA1* or *BRCA2* pathogenic variants (g*BRCA1/2* PV).^1–4^ g*BRCA1/2* PV carriers have an elevated lifetime breast cancer risk of 60-80%.^5–9^ The UK Prospective study of Outcomes in Sporadic versus Hereditary breast cancer (POSH) demonstrated similar overall survival (OS) between young-onset female breast cancer patients with and without g*BRCA1/2* PVs.^3^

*BRCA1* and *BRCA2* are critical for repairing double-stranded DNA breaks through homologous recombination, and complete loss of *BRCA1/2* function through allele-specific loss of heterozygosity (asLOH) is associated with homologous recombination deficiency (HRD).^10–12^ Although both proteins function in the same pathway, some broad differences in somatic features between g*BRCA1* and g*BRCA2* tumors have been reported. g*BRCA1* tumors are mostly ER-negative, whereas g*BRCA2* tumors are mostly ER-positive.^13,14^ Compared to g*BRCA2* tumors, g*BRCA1* tumors harbor high frequencies of *TP53* PVs and greater amounts and distinct patterns of copy number variants (CNV).^15–20^ Losses of chr2q, chr4p, chr4q, chr5q, and chr12q are frequent in g*BRCA1*-associated tumors, whereas losses of chr6q and chr13q occur in g*BRCA2*-associated tumors.^19–21^ Furthermore, g*BRCA1*-associated tumors exhibit greater activation of genes involved in DNA repair than g*BRCA2*-associated tumors.^22^ However, the full landscape of differences between g*BRCA1* and g*BRCA2* tumors remains incomplete, as previous studies have been limited by low numbers of carriers and by the use of older technologies such as comparative genome hybridization. Whether molecular differences between g*BRCA1* and g*BRCA2* tumors have therapeutic utility has not been elucidated.

g*BRCA1/2-*associated tumors are sensitive to poly (ADP-ribose) polymerase inhibitors (PARPi) in both metastatic and high-risk early breast cancers.^23–27^ Adjuvant PARPi significantly improves overall survival (OS) in early-stage, high-risk disease^28^ and improves progression-free survival in metastatic disease.^25,26^ In ER-positive, HER2-negative tumors, adjuvant cyclin-dependent kinase 4/6 inhibitors (CDK4/6i) combined with endocrine therapy improve invasive disease-free survival in early-stage disease, and significantly increase OS in metastatic disease, compared to endocrine therapy alone.^29–32^ Thus, both PARPi and CDK4/6i confer clinically meaningful benefit in early and metastatic breast cancer.

Despite the efficacy of both PARPi and CDK4/6i, primary and secondary resistance to each therapy has been observed.^26,28,33–38^ PARPi resistance most commonly arises through reversion mutations of g*BRCA1/2.*^39^ Absence of asLOH^35^ also may confer primary resistance to PARPi in g*BRCA1/2* PV carriers.^12,40^ We previously reported differences in asLOH frequency between g*BRCA1* and g*BRCA2* breast cancers from PV carriers unselected for age (90% vs 54%).^12^ CDK4/6i impede cell proliferation by inhibiting the phosphorylation of the retinoblastoma protein (Rb) during the cell cycle.^41,42^ Genomic alterations, such as *RB1* loss and *MYC* amplification, have been implicated in resistance to CDK4/6i.^43,44^

In young-onset g*BRCA1/2*-associated breast cancer, the frequency of asLOH and its impact on survival remain unclear, as does the landscape of genomic alterations associated with response to CDK4/6i. An improved understanding of the distinct genomic landscapes of g*BRCA1* and g*BRCA2* PV-associated breast cancer may inform current treatment selection, particularly in ER-positive, HER2-negative tumors where both PARPi and CDK4/6i are treatment options, and may inform strategies for possible future therapy development. Thus, we performed comprehensive genomic profiling of treatment-naïve, primary non-metastatic breast tumors using whole exome sequencing (WES) of matched tumor-germline DNA in 136 young-onset breast cancer patients with g*BRCA1/2* PVs from the POSH cohort.

## Methods

### Study description

The POSH prospective cohort study (UK, 2000-2008) recruited over 3,000 women with invasive breast cancer at age ≤40 years to investigate the impact of g*BRCA1/2* PVs on breast cancer outcomes.^45^ The primary outcome analysis was published in 2018 with a median follow-up of 8.2 years.^3^ Written informed consent was obtained for all participants at recruitment for further analysis of their tissue and clinical data; ethical approval was granted in 2000 (MREC00/6/69).

Participants with g*BRCA1/2* PVs were identified using sequencing and multiplex ligation probe analysis (MLPA) in patients meeting UK thresholds for clinical germline genetic testing at the time.^3,46^ Three hundred and ninety-one patients with g*BRCA1/2* PVs were identified: 55 patients were excluded as they received neoadjuvant chemotherapy. An additional 135 patients were excluded because tumor blocks were unavailable.

### Sequencing and bioinformatic analysis

All H&Es were reviewed to ensure breast cancer diagnosis by a board-certified pathologist (AN). Following exclusion of patient samples with insufficient tumor as judged by AN, failed DNA extraction or failed library preparation, and those without tumor-germline matches, we performed WES of 160 formalin-fixed paraffin-embedded (FFPE) tumor blocks and 194 germline DNAs (Figure S1). After tumor-germline matching, removal of duplicate tumor samples, and exclusion of three tumors that were metastatic at diagnosis, two tumors from patients without g*BRCA1/2* PVs, and one tumor sample with poor sequencing coverage, we had 136 matched tumor-germline sequenced samples (Figure S1). The only significant differences between the 136 and the full set of 391 g*BRCA1/2* PV carriers from POSH were receipt of neoadjuvant chemotherapy, pathological M-stage, and unknown pathological T-stage, all of which were due to pre-determined study eligibility criteria (Table S1). The eligibility for the overall study included women diagnosed with breast cancer between ages 41-50 years if known g*BRCA1/2* positive. We called and filtered somatic single nucleotide variants (SNVs), indels, and copy number variants (CNVs) in 598 cancer genes (Table S2) using a custom computational pipeline (Figure S2).

We selected 81 women with ER-positive, HER2-negative invasive breast cancer diagnosed before age 50 years without g*BRCA1/2* PVs^12^ in The Cancer Genome Atlas (TCGA) for use as noncarrier comparators. We retrieved BAM files from the NCI Genomic Data Commons^47^, excluding six patients with germline PVs in *CHEK2*, *LZTR1, MSH6*, *PALB2*, and *POT1.* Somatic SNVs, indels, and CNVs were called using the same custom computational pipeline used for POSH samples in 66 unique tumor-germline samples (Figures S2, S3).

Details of asLOH determination, HRD-score calculation, single-base substitution (SBS) signature fitting, and TMB calculation are in Supplemental Methods.

### Statistical analysis

We performed Firth logistic regression to determine the enrichment of chromosomal arm CNVs and alterations in genes by germline variant gene, ER-status, and HRD status (threshold HRD-high ≥ 42) in R.^48^ Analyses were adjusted for ER-status, germline variant gene, asLOH status, and multiple testing as appropriate. We compared mean HRD scores using two-sided t-tests and ANOVA, and compared median SBS proportions and TMB scores using Mann-Whitney and Kruskal-Wallis nonparametric tests. Survival analysis was performed using the Kaplan-Meier survival estimator; we estimated hazard ratios and 95% confidence intervals (CIs) using the Mantel-Haenszel method and estimated p-values using the Log-rank (Mantel-Cox) test. Survival analyses were performed in GraphPad Prism v10.

We assigned genes to pathways based on Hallmark Pathway^49^ classification and performed enrichment analysis using Firth logistic regression, as above, with the Hallmark Pathways as dependent variables. Analyses were adjusted for ER-status, germline variant gene, and asLOH status as appropriate.

## Results

### Cohort description

We evaluated WES results of 136 treatment-naïve matched tumor-germline samples from g*BRCA1/2* positive women diagnosed with young-onset breast cancer participating in the POSH study (Figure S1). Eighty-six (63.2%) and 50 (36.8%) women had g*BRCA1* and g*BRCA2* germline PVs, respectively. The median age at diagnosis was 36 years, and 92.6% of women were ≤ age 40 (Table 1). Overall, 65 (47.8%) women had ER-positive tumors. Demographic, tumor pathology, and treatment data are shown in Table 1.

**Table 1:** Characteristics of study participants.

| Characteristics of study participants, N (%) |  |  |  |  |
| --- | --- | --- | --- | --- |
|  | Total | BRCA1 | BRCA2 | P-value |
|  | 136 (100) | 86 (63.2) | 50 (36.8) |  |
| **Age** |  |  |  |  |
| 18-25 | 3 (2.2) | 2 (2.3) | 1 (2.0) | 1 |
| 26-30 | 13 (9.6) | 11 (12.8) | 2 (4.0) | 0.132 |
| 31-35 | 41 (30.1) | 30 (34.9) | 11 (22.0) | 0.126 |
| 36-40 | 69 (50.7) | 36 (41.9) | 33 (66.0) | 0.008 |
| 41-50 | 10 (7.4) | 7 (8.1) | 3 (6.0) | 0.745 |
| **Ethnicity** |  |  |  |  |
| White/Caucasian | 129 (94.9) | 82 (95.3) | 47 (94.0) | 0.708 |
| Black | 4 (2.9) | 3 (3.5) | 1 (2.0) | 1 |
| Asian | 2 (1.5) | 1 (1.2) | 1 (2.0) | 1 |
| Unknown | 1 (0.7) | 0 (0.0) | 1 (2.0) | 0.368 |
| **Histological subtype** |  |  |  |  |
| Ductal | 122 (89.7) | 79 (91.9) | 43 (86.0) | 0.381 |
| Lobular | 6 (4.4) | 0 | 6 (12.0) | 0.002 |
| Medullary | 4 (2.9) | 4 (4.7) | 0 | 0.296 |
| Mixed | 3 (2.2) | 2 (2.3) | 1 (2.0) | 1 |
| Unknown | 1 (0.7) | 1 (1.2) | 0 | 1 |
| **Histological grade** |  |  |  |  |
| 1 | 1 (0.7) | 1 (1.2) | 0 | 1 |
| 2 | 19 (14.0) | 4 (4.7) | 15 (30.0) | <0.001 |
| 3 | 115 (84.6) | 80 (93.0) | 35 (70.0) | 0.001 |
| Unknown | 1 (0.7) | 1 (1.2) | 0 | 1 |
| **Hormone receptor status** |  |  |  |  |
| ER |  |  |  |  |
| Negative | 71 (52.2) | 67 (77.9) | 4 (8.0) | <0.001 |
| Positive | 65 (47.8) | 19 (22.1) | 46 (92.0) | <0.001 |
| PR |  |  |  |  |
| Negative | 65 (47.8) | 58 (67.4) | 7 (14.0) | <0.001 |
| Positive | 41 (30.1) | 10 (11.6) | 31 (62.0) | <0.001 |
| Unknown | 30 (22.1) | 18 (20.9) | 12 (24.0) | 0.674 |
| HER2 |  |  |  |  |
| Negative | 107 (78.7) | 66 (76.7) | 41 (82.0) | 0.522 |
| Positive | 11 (8.1) | 4 (4.7) | 7 (14.0) | 0.098 |
| Unknown | 18 (13.2) | 16 (18.6) | 2 (4.0) | 0.017 |
| **Pathological T stage** |  |  |  |  |
| 1 | 68 (50.0) | 43 (50.0) | 25 (50.0) | 1 |
| 2 | 61 (44.9) | 39 (45.3) | 22 (44.0) | 1 |
| 3 | 7 (5.1) | 4 (4.7) | 3 (6.0) | 0.708 |
| **Pathological N stage** |  |  |  |  |
| 0 | 69 (50.7) | 54 (62.8) | 15 (30.0) | <0.001 |
| 1 | 66 (48.5) | 31 (36.0) | 35 (70.0) | <0.001 |
| Unknown | 1 (0.7) | 1 (1.2) | 0 | 1 |
| **Treatment** |  |  |  |  |
| Chemotherapy regimen |  |  |  |  |
| Anthracyclines | 103 (75.7) | 65 (75.6) | 38 (76.0) | 1 |
| Anthracyclines + Taxanes | 26 (19.1) | 16 (18.6) | 10 (20.0) | 0.825 |
| Other | 2 (1.5) | 2 (2.3) | 0 | 0.532 |
| None | 5 (3.7) | 3 (3.5) | 2 (4.0) | 1 |
| Chemotherapy period |  |  |  |  |
| Adjuvant | 131 (96.3) | 83 (96.5) | 48 (96.0) | 1 |
| None | 5 (3.7) | 3 (3.5) | 2 (4.0) | 1 |
| Definitive surgery |  |  |  |  |
| Breast-conserving surgery (BCS) | 72 (52.9) | 49 (57.0) | 23 (46.0) | 0.285 |
| Mastectomy | 63 (46.3) | 36 (41.9) | 27 (54.0) | 0.212 |
| Nodal surgery only | 1 (0.7) | 1 (1.2) | 0 | 1 |

### Molecular features in breast cancers

We evaluated the genomic landscape of the breast cancers, including asLOH, HRD, TMB, and SBS. Tumors had high rates of asLOH when stratified by gene: g*BRCA1* (93%) and g*BRCA2* (96%), and by ER-status: ER-negative (94.4%) and ER-positive (93.8%) (Table S3). HRD scores were significantly higher in tumors with asLOH compared to tumors without asLOH (nonLOH) in both g*BRCA1* (57.4 ± 1.3 vs 22.6 ± 6.1, p<0.0001) and g*BRCA2* (43.7 ± 1.5 vs 23.5 ± 6.5, p=0.005) PV carriers, and when grouped by ER-status: ER-negative (57.6 ± 1.3 vs 22.8 ± 9.2, p<0.0001) and ER-positive tumors (46.4 ± 1.6 vs 22.8 ± 4.1, p<0.001) (Figure 1a, Table S3). g*BRCA1* tumors with asLOH had significantly higher HRD scores than g*BRCA2* tumors with asLOH (57.4 ± 1.3 vs 43.7 ± 1.5, p<0.0001) (Figure 1a).

**Figure 1:**
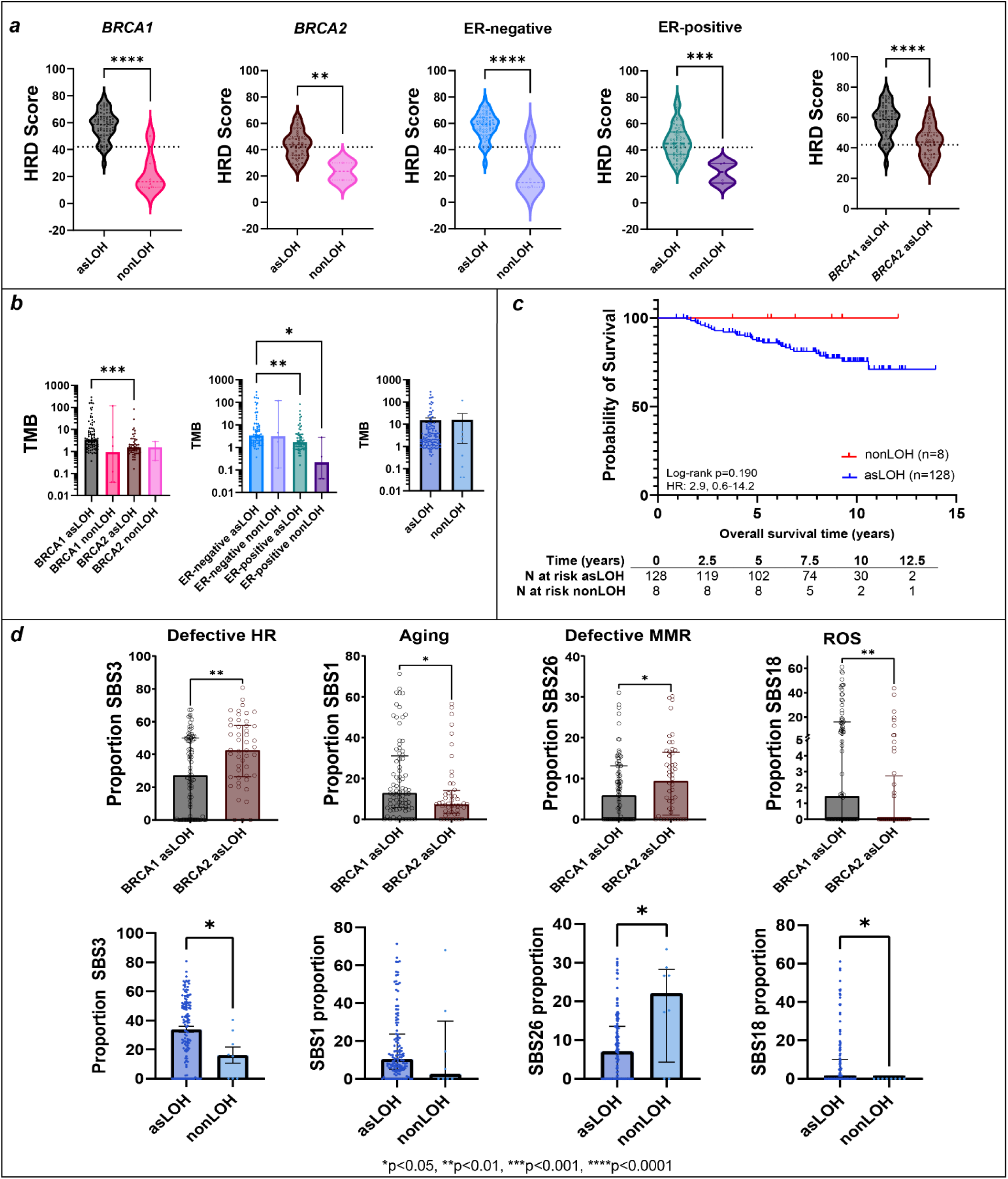
Molecular features of breast cancer in *BRCA1* and *BRCA2* pathogenic variant carriers with overall survival. a) Homologous recombination deficiency (HRD) scores in tumors with allele-specific loss of heterozygosity (asLOH) compared to those without, stratified by germline variant gene and ER-status. Dotted black line is HRD score = 42. b) Median tumor mutational burden (TMB) by asLOH status, germline gene variant, and ER-status. c) Overall survival by asLOH. d) Median of the proportion of single base substitution signatures 3 (SBS3), SBS1, SBS26, and SBS18 in tumors asLOH by gene and in tumors with asLOH and those without. SEM = standard error of mean, IQR = interquartile range.

Tumors with g*BRCA1* asLOH had higher median TMB than those with g*BRCA2* asLOH (3.4, IQR 1.6-11.2 vs 1.5, IQR 0.9-3.4, p=0.001). ER-positive tumors with asLOH had significantly lower median TMB than ER-negative tumors with asLOH (1.7, IQR 0.9-3.9 vs 3.4, IQR 1.6-11.0, p=0.008) (Figure 1b). However, there was no difference in median TMB between tumors with and without asLOH.

We evaluated breast cancer overall survival (OS) by tumor molecular features. Survival in women with nonLOH tumors was 100% throughout the 8.2-year median follow-up period, but not statistically significantly different from that of women with tumors with asLOH (Figure 1c). Favorable survival for women with nonLOH tumors remained when the analysis was stratified by germline variant gene or by ER-status (Figure S4). Although not significant, women with HRD high (≥42) tumors tended to do worse (Figure S5).

SBS3, the signature associated with g*BRCA1*/2 PVs,^35^ was the most abundant signature in all tumors with an average contribution of 32.8% (Figure S6). SBS1 (aging-associated), SBS26 (mismatch repair deficiency-associated), and SBS18 (reactive oxygen species (ROS)-associated) were the next most abundant signatures.^35^ g*BRCA1* tumors had significantly higher median proportional contribution of SBS1 (12.9 vs 7.3, p=0.013) and SBS18 (1.4 vs 0, p=0.007), and significantly lower proportions of SBS3 (27.3 vs 42.6, p=0.002) and SBS26 (5.9 vs 9.4, p=0.049) than g*BRCA2* tumors. The median proportional contributions of SBS3 and SBS18 in tumors with asLOH were significantly higher than in nonLOH tumors (SBS3 38.5 vs 15.9, p=0.023; SBS18 0 vs 0, p=0.023) (Figure 1d, Table S4).

### Copy number variation in tumors

Copy number analysis revealed chromosome arm chr1q and chr8q gains, both prevalent in breast cancers,^50^ in all tumors (Figure 2a). Eighty-eight and 108 tumors had single copy loss of *RB1* and *TP53*, respectively. Copy loss of *RB1* and *BRCA2,* both located on chromosome 13, occurred on the same segment significantly more frequently than expected by chance (56/88, p =0.010), whereas copy loss of *TP53* and *BRCA1,* both on chromosome 17, occurred on the same segment significantly less frequently than expected by chance (22/108, p<0.001) (Figure 2a). Thus, it appears that *RB1* and *BRCA2* loss are non-independent, whereas copy number loss of *TP53* and *BRCA1* are independent of each other. The loss of *RB1* (*RB1* asLOH) occurred significantly more frequently in g*BRCA2* asLOH tumors than g*BRCA1* asLOH tumors, however, there was no difference in the frequency of *RB1* PVs or *RB1* biallelic loss. Conversely, *TP53* asLOH was not different between g*BRCA1* asLOH and g*BRCA2* asLOH tumors, although g*BRCA1* asLOH tumors had significantly more *TP53* PVs and *TP53* biallelic loss than g*BRCA2* asLOH tumors (Figure 2a).

**Figure 2:**
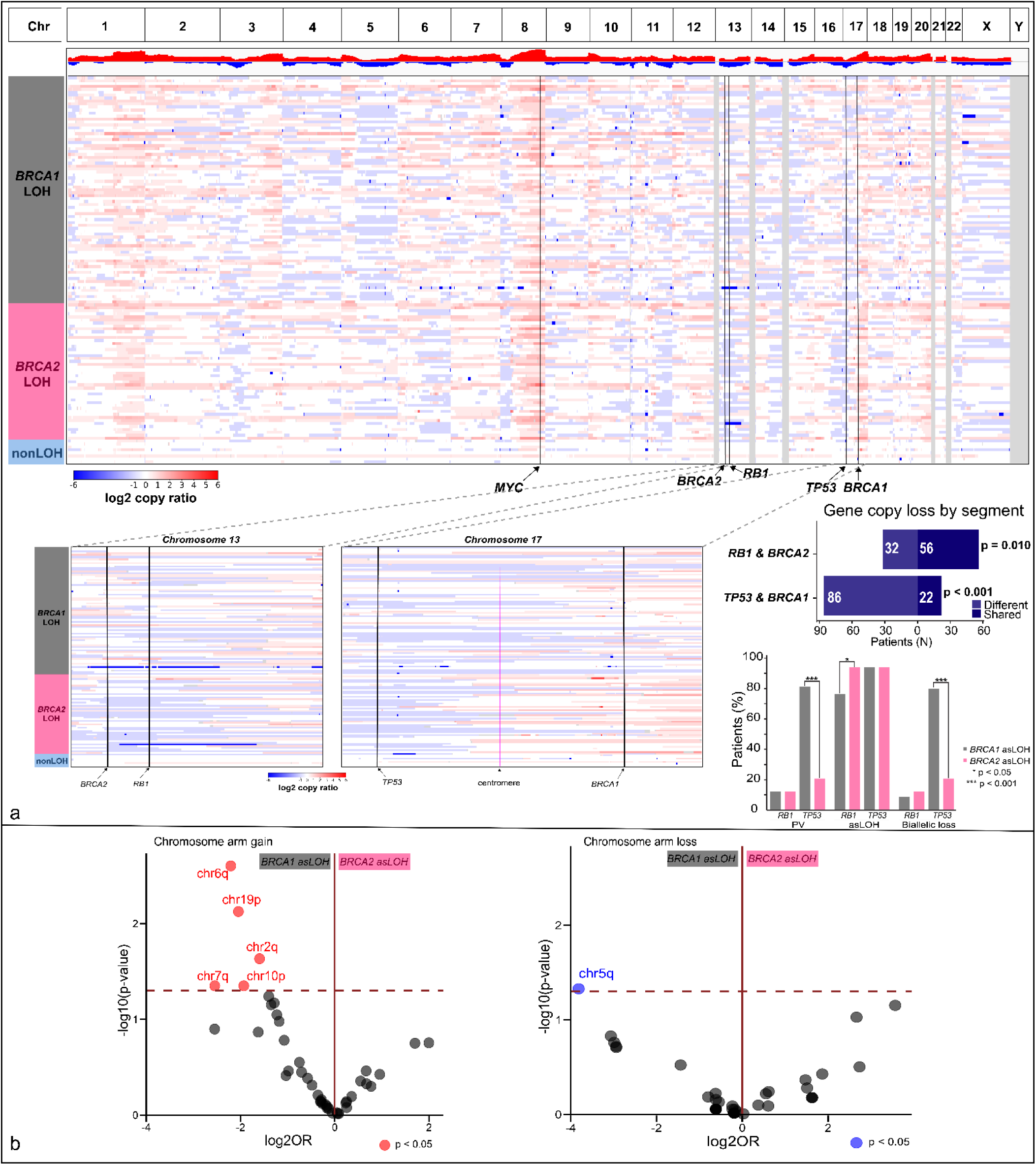
Copy number variation in tumor samples. a) Landscape of copy number variants in tumors. Insert shows zoomed in images of chromosomes 13 and 17 and the quantified values of times that *RB1* shares a copy number loss segment with *BRCA2* and *TP53* shares one with *BRCA1*. Verticle bar graph shows the frequency of *RB1* and *TP53* pathogenic variants (PV), allele-specific loss of heterozygosity (asLOH), and biallelic loss in *BRCA1* and *BRCA2* tumors with asLOH. b) Enrichment of chromosome arm gains and losses in tumors with *BRCA1* asLOH compared to those with *BRCA2* asLOH adjusted for estrogen receptor (ER)-status. Enrichment was performed using Firth logistic regression. Dashed line is -log10p-value = 1.3.

We observed differences in chromosome arm copy number between tumors with g*BRCA1* asLOH and g*BRCA2* asLOH: gains of chr1q, chr2q, chr6q, chr6p, chr9p, chr10p, chr11q, chr12p, and chr19p were significantly enriched in tumors with g*BRCA1* asLOH, whereas copy number loss of chr6q was significantly enriched in tumors with g*BRCA2* asLOH (Figure S7). As gains in chr1q, chr10p, and chr12p are frequent in ER-negative tumors,^51^ we adjusted the analysis for ER-status, and found that chr6q, chr19p, chr2q, chr7q, and chr10p gains, and chr5q copy number loss remained enriched significantly in tumors with g*BRCA1* asLOH compared to those with g*BRCA2* asLOH and chr6q loss was no longer enriched in tumors with g*BRCA2* asLOH (Figure 2b).

### Enrichment of gene-level alterations

To further define the complete genomic landscape of the tumors, we assessed gene-level somatic alterations (pathogenic SNVs/indels and CNVs) in 598 cancer-associated genes (Table S2). In the 136 tumors evaluated, we found 4146 pathogenic SNV/indels and 4205 pathogenic CNVs, in 572/598 genes (Table S5). Pathogenic SNV/indels in the 50 most commonly altered genes are shown in Figure S8.

When we compared pathogenic SNV/indels and CNVs in tumors from g*BRCA1* PV carriers to those from g*BRCA2* PV carriers, with adjustment for ER-status, we observed a larger number of somatic alterations significantly enriched in g*BRCA1* tumors (Figure 3a, Table S6). Pathway analysis revealed enrichment of pathogenic SNV/indels/CNVs in genes in multiple Hallmark pathways,^49^ including the ROS pathway, DNA repair, and epithelial-mesenchymal transition in g*BRCA1* tumors compared to g*BRCA2* tumors (Figure 3a).

**Figure 3:**
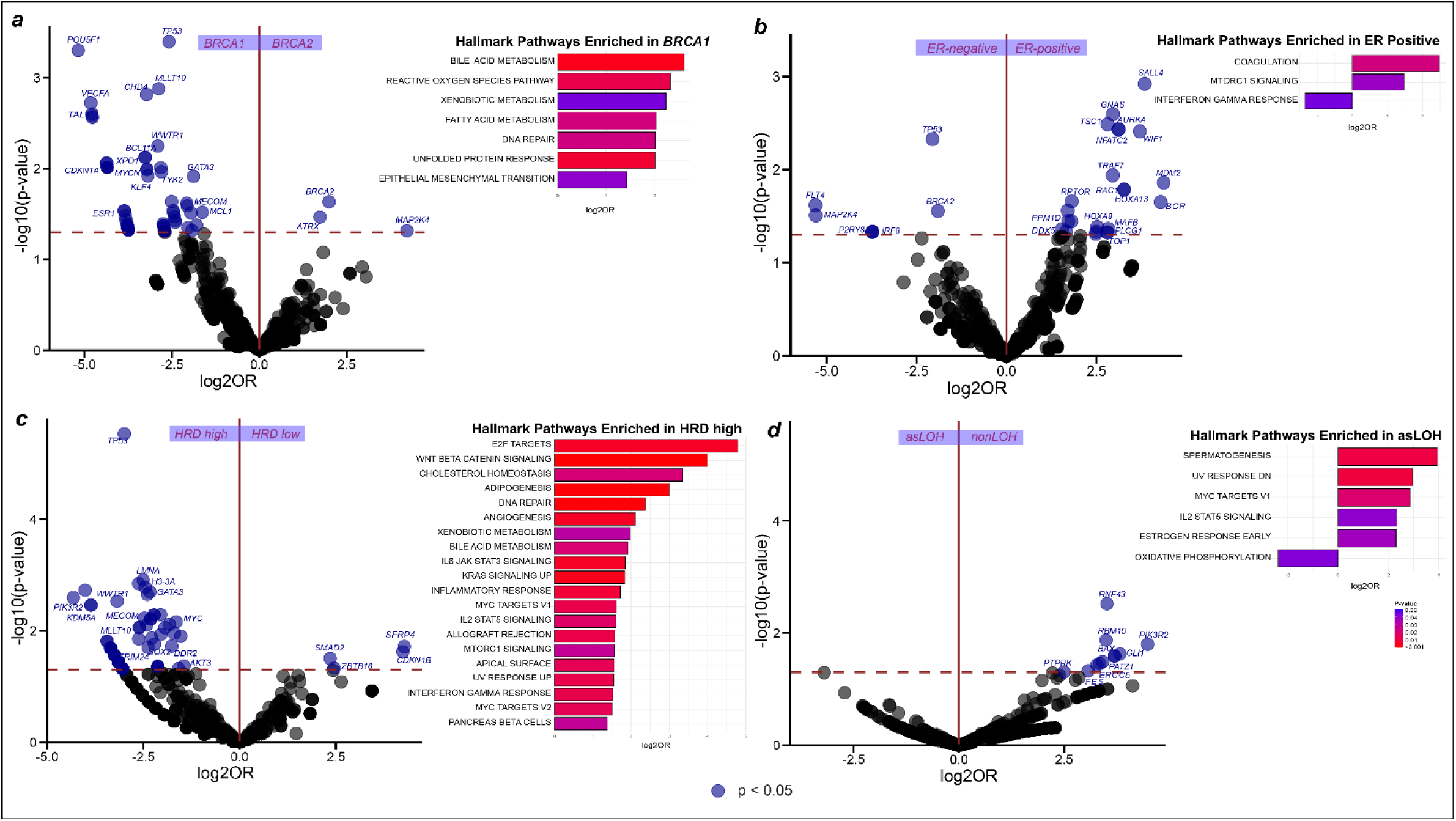
Differential enrichment of somatic variants, inclusive of pathogenic single nucleotide variants, indels, and copy number alterations, and Hallmark Pathway enrichment in breast cancers. Enrichment of somatic variants and enrichment of Hallmark Pathway alterations by a) germline variant gene adjusted for estrogen receptor (ER)-status; b) by ER-status adjusted for germline variant gene; c) by homologous recombination deficiency (high ≥ 42) adjusted for allele-specific loss of heterozygosity (asLOH); and d) by asLOH stratified by germline gene variant. Enrichment analysis was performed using Firth logistic regression. Dashed line is -log10p-value = 1.3.

With adjustment for g*BRCA1/2* PV carrier status, ER-positive tumors had greater enrichment of pathogenic somatic SNV/indel/CNV variants compared to ER-negative tumors (Figure 3b, Table S7) and were enriched for alterations in genes in the coagulation and MTORC1 signaling Hallmark pathways. ER-negative tumors were enriched for alterations in the interferon gamma response pathway (Figure 3b). The total number of genes with SNV/indels/CNV and SNV/indels was higher in ER-negative tumors compared to ER-positive tumors when the analysis was performed without adjustment for g*BRCA1/2* PV status, as seen in previous studies (Figure S9).^52^

More genes were enriched in SNV/indels and CNVs in HRD-high tumors compared to HRD-low tumors, adjusted for LOH status (Figure 3c, Table S8). Genes in the E2F targets and WNT/Beta-Catenin signaling pathways were the most highly enriched in HRD-high tumors (Figure 3c). Tumors without asLOH were enriched for alterations in the oxidative phosphorylation pathway compared to those with asLOH (Figure 3d, Table S9).

### Presence of CDK4/6 resistance alterations

Genetic alterations implicated in acquired and intrinsic resistance include alterations in *RB1, AURKA, MYC, CCNE1, SPEN, FAT1, ARID1A, PTEN, FGFR1,* and *EGFR*.^38,53–58^ We evaluated the presence of somatic alterations (SNVs/indels, CNVs-including single copy loss) implicated in resistance to CDK4/6i in ER-positive, HER2-negative tumors within the POSH cohort and found that all tumors contained at least one alteration implicated in resistance (Figure 4).^56^ To determine whether g*BRCA1/2* carriers were more likely than noncarriers to have somatic alterations in CDK4/6i resistance-associated genes, we compared rates of alteration between carriers from POSH and noncarriers from the TCGA, all with ER-positive, HER2-negative tumors. To mitigate any potential bias in sequencing coverage across the two cohorts, we limited our analysis to genes with at least one alteration detected in the TCGA cohort (Table S2). We found statistically significant enrichment of alterations in several genes in tumors from the POSH cohort compared to those from the TCGA, including *RB1* (OR:6.3, 95% CI:2.8-15.4, p_adj_=0.001), *TP53* (OR:4.6, 95% CI:1.9-12.1, p_adj_=0.017), *FAT1* (OR:3.9, 95%CI:1.84-8.7, p_adj_=0.013), and *MYC* (OR:4.0, 95%CI:1.8-9.1, p_adj_=0.017), all of which are associated with resistance to CDK4/6i (Figure 4).

**Figure 4:**
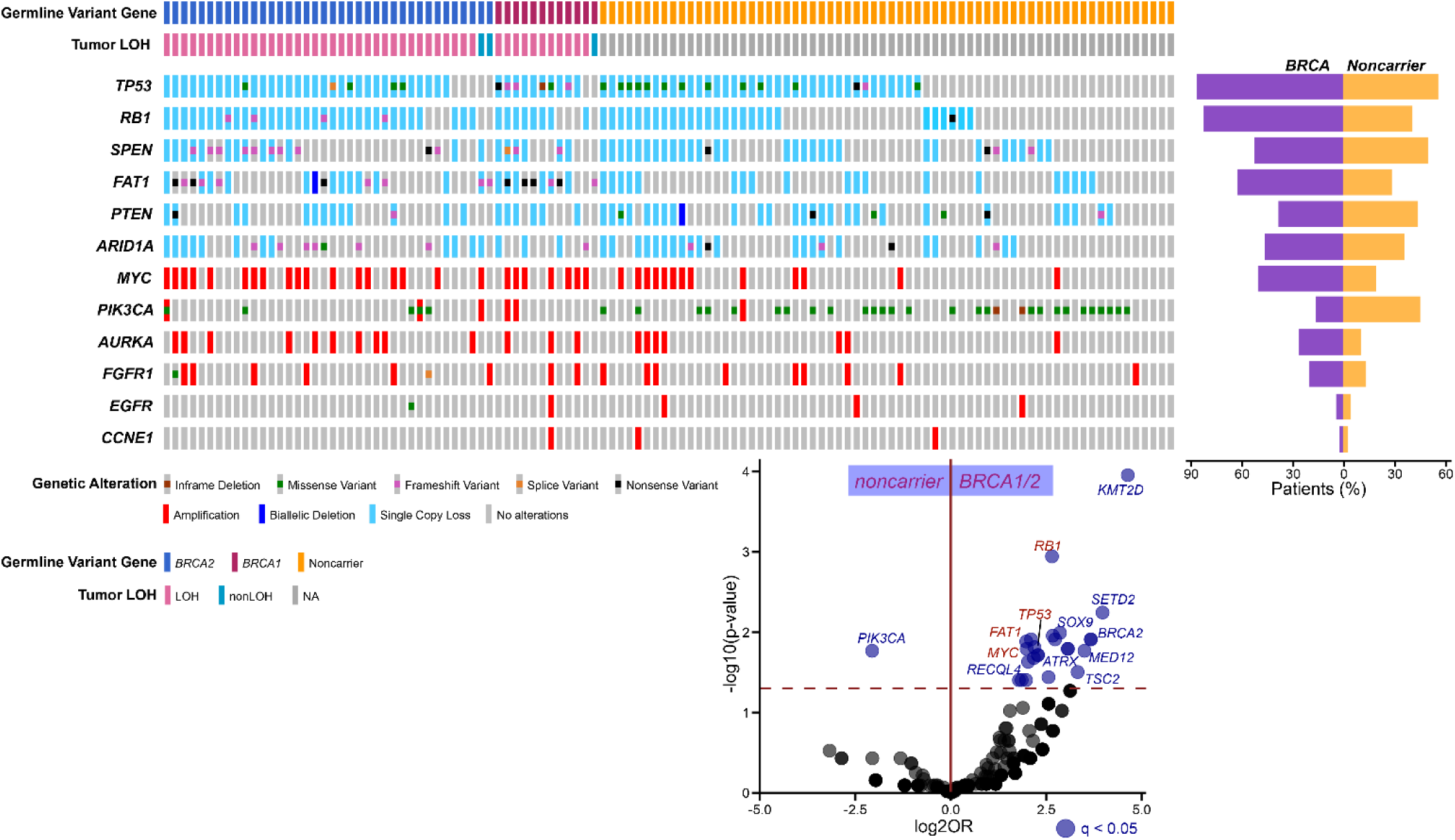
Enrichment of somatic variants associated with resistance to CDK4/6 inhibitors in ER-positive, HER2-negative tumors from *BRCA1/2* carriers in POSH compared to noncarriers from the TCGA. Oncoprint of somatic variants in tumors, frequency of somatic variants, and enrichment of variants in POSH vs TCGA samples. Dashed line is -log10p-value = 1.3, q is the false discovery rate (FDR)-adjusted p-value.

## Discussion

We performed WES on treatment-naïve matched tumor-germlines from 136 young g*BRCA1/2* PV carriers from the POSH cohort and found high levels of asLOH, over 90%, in all tumors regardless of germline variant gene and tumor ER-status. No deaths were seen in women with a nonLOH breast cancer over the 8.2-year follow-up period; however, these numbers were small, and survival differences with asLOH tumors were not statistically significant. We observed numerous significant differences between the genomic landscape of g*BRCA1* and g*BRCA2* PV-associated breast cancers, with implications for future therapeutic selection. When evaluating ER-positive, HER2-negative breast cancers, we found significant enrichment of genetic changes associated with CDK4/6i resistance mechanisms.

Comparing g*BRCA1* and g*BRCA2* breast cancers with asLOH, g*BRCA1* tumors had significantly higher TMB score (although the median TMB was low at 3.4), HRD scores, and proportions of aging-associated SBS1 and ROS-associated SBS18. g*BRCA2* asLOH tumors had higher proportions of SBS3 and SBS26 which are associated with defective DNA repair through homologous recombination and mismatch repair, respectively.^35,59^ g*BRCA1* tumors, compared to g*BRCA2* tumors, had enrichment of multiple chromosome arm gains and amplifications with chr6q most significant even after correction for ER-status. Others also have found greater numbers of CNVs in g*BRCA1* than g*BRCA2* tumors, focal gains and amplifications of chr6q in g*BRCA1* tumors, and deletions of chr6q in g*BRCA2* tumors.^20,21,60,61^ We show that the deletions of chr6q which are significantly enriched in g*BRCA2* tumors are dependent on ER-status, as the association is lost with adjustment for ER-status, and that chr6q gains are g*BRCA1* specific. Chr6q contains cancer-associated genes *ROS1*,^62^ *FOXO3*, which, in addition to promoting breast cancer growth and metastasis, regulates ROS,^63,64^ and *RSPO3*, which drives the formation of hormone receptor-negative breast cancer and is associated with EMT.^65^ We find significantly more copy number gains of *ROS1*, *FOXO3*, *RSPO3*, and significantly more *FOXO3* amplifications in g*BRCA1* compared to g*BRCA2* tumors. Compared to g*BRCA2* tumors, g*BRCA1* tumors were enriched for alterations in ROS, EMT, and DNA repair pathways. Loss of *BRCA1* has been shown to promote EMT, increase ROS, and dysregulate DNA repair pathways in other studies.^22,66–68^ The observed molecular differences suggest that g*BRCA1* and g*BRCA2* tumors may respond differently to molecularly targeted therapies such as ROS1 inhibitors^62,69^ and anti-RSPO3 antibodies,^70,71^ which are under investigation for cancer treatment in the preclinical and clinical settings.

It has been suggested that *RB1* asLOH occurs simultaneously with *BRCA2* asLOH in tumors with g*BRCA2* PVs due to the proximity of the genes on chr13.^56,72,73^ Inagaki-Kawata et al. suggested also that *TP53* loss on chr17 cooccurred with *BRCA1* loss, but did not determine whether the loss occurred on the same or different segments.^72^ We found that *RB1* asLOH was more frequent in g*BRCA2* tumors than g*BRCA1* tumors and cooccurred on CNV segments with *BRCA2* asLOH. There was no difference in the frequency of *TP53* asLOH between g*BRCA1* and g*BRCA2* tumors; the frequency of *TP53* asLOH was greater than 90% in both tumor types and *TP53* asLOH was rarely on the same segments as *BRCA1* asLOH. A previous study suggested that during g*BRCA1*-associated breast cancer development, *TP53* asLOH occurred before *BRCA1* loss.^74^ Thus, *RB1* asLOH is dependent on *BRCA2* asLOH but *TP53* asLOH is independent of *BRCA1* asLOH in breast cancer.

The availability of FDA-approved targeted therapies for the treatment of g*BRCA1/2* driven tumors and for ER-positive, HER2-negative tumors presents a treatment choice decision when managing g*BRCA1/2* carriers with ER-positive, HER2-negative tumors.^25,26,29–32^ Previous studies of breast tumors unselected for age at diagnosis showed levels of asLOH as low as 54% in g*BRCA2* PV carriers.^12^ In this study of young women, we found high levels of asLOH, greater than 90%, in both g*BRCA1* and g*BRCA2* PV carriers, regardless of tumor ER-status. This finding suggests that most breast cancers in young women with g*BRCA1/2* PVs may be responsive to PARP inhibition. It is likely that in populations with older g*BRCA1/2* carriers, sporadic tumors are mixed in with g*BRCA1/2*-associated breast cancers, but young patients have mostly g*BRCA1/2*-associated tumors. Two retrospective studies in patients with advanced breast cancer have suggested g*BRCA2* PV carriers with advanced breast cancer have attenuated responses to CDK4/6i plus endocrine therapy.^75,76^ Bruno et al. reported lower progression-free and overall survival in carriers of g*BRCA1/2*, g*ATM*, and g*CHEK2* PVs compared to noncarriers, and Kim et al. identified g*BRCA2* PVs as associated with lower progression-free survival following treatment with CDK4/6i, albeit with only five g*BRCA2* PV carriers. ^75,76^ A larger study in patients with metastatic breast cancer similarly reported lower progression-free survival rates for g*BRCA2* PV carriers treated with CDK4/6i compared to noncarriers.^56^ Our findings in young women with early breast cancer suggest genomic mechanisms that underline resistance to CDK4/6 inhibitors, including SNV/indels and CNVs in *RB1*, *TP53*, *FAT1*, and *MYC*. Thus, for young women with g*BRCA1/2* PVs and ER-positive, HER2-negative breast cancer, PARPi, rather than CDK4/6i, may be the optimal choice in the adjuvant and metastatic settings when both options are being considered. An ongoing clinical trial (NCT06380751) is testing PARPi versus CDK4/6i in g*BRCA1/2* PV carriers with metastatic breast cancer.

The main limitation was the low number of participants with nonLOH tumors, which may have limited our ability to detect significant survival differences. We evaluated only pretreatment samples and thus can only make predictions about potential treatment responses that will need to be tested in subsequent studies.

In conclusion, we show that tumors from young g*BRCA1/2* PV carriers with breast cancer almost always have asLOH. We also show differences between the molecular features of g*BRCA1* and g*BRCA2* tumors which may impact the suitability of molecularly targeted therapies. Importantly, we show enrichment of gene alterations associated with resistance to CDK4/6 inhibition in ER-positive, HER2-negative tumors from g*BRCA1/2* PV carriers compared to noncarriers. Given the high levels of asLOH and presence of CDK4/6i resistance-associated alterations, our data suggest that PARPi may be preferable over CDK4/6i in young g*BRCA1/2* carriers with ER-positive, HER2-negative breast cancer when both therapies are considered in the adjuvant and metastatic setting.

## Data Availability

All data produced in the present study are available upon reasonable request to the authors

## Data Supplement

### Supplemental Methods

#### Whole-exome sequencing (WES) of tumor and germline DNA

Germline DNA libraries were prepared using the NEBNext Ultra DNA Library Prep Kit (New England Biolabs). Tumor DNA was extracted from FFPE using standard laboratory deparaffinization, proteinase K digestion, and ethanol precipitation. Tumor DNA libraries were prepared using the NEBNext FFPE DNA Repair Mix and NEBNext Ultra II DNA Library Prep Kit (New England Biolabs). DNA libraries were pooled and hybridized using SureSelect Human All Exon v7 for Illumina Multiplexed Sequencing (Agilent). All DNA samples, libraries, and hybridization pools were quantified with Qubit 2.0 Fluorometer (ThermoFisher), and fragment size was determined by BioAnalyzer 2100 (Agilent). WES was performed with 150 paired end reads on Illumina NovaSeq 6000 by the University of Pennsylvania Next Generation Sequencing Core.

#### Bioinformatics Analysis: Data Preprocessing

Tumor and germline BAM files were aligned to the GRCh38 reference genome using Burrows-Wheeler Aligner (BWA v.0.7.17).^1^ Sequencing quality metrics were generated by Samtools (v.1.20).^2^ Pathogenic germline variants in *BRCA1/2* were called by DeepVariant (v.1.6.0) and VarDict and were confirmed with prior clinical sequencing records for each patient at the University of Southampton.^3,4^ We implemented an “identity by descent” (IBD) algorithm (PLINK v.1.9), using common biallelic SNV (SNP) locations to confirm matching of tumor and germline DNA samples with IBD scores > 0.9.^5,6^

#### Bioinformatics Analysis: Identification of Somatic Pathogenic SNV and Indels

Somatic single-nucleotide variants (SNV) and indels were called and filtered using a custom computational pipeline (Figure S2). SNVs and indels were first called by a combination of Lancet (v.1.1.0), Mutect2 (v.4.2.0.0), Strelka2 (v.2.9.2), VarDict, and Varscan2 (v.2.4.4), and then annotated using Ensembl Variant Effect Predictor (VEP v.112).^4,7–11^ All somatic variants were preprocessed using Varlociraptor (v.8.4.6) and custom scripts to remove potential sequencing and FFPE artifacts.^12^ We selected variants with Varlociraptor somatic variant probability ≥ 0.9 in cancer genes (Table S2) for inclusion in our analysis for POSH but not TCGA samples because the lower sequencing depth in the TCGA samples skewed the Varlociraptor probability calls.^13–15^ We cleaned variants to remove those we determined as duplicates, such as SNVs called by Strelka2 and Varscan, which overlapped with multi-nucleotide variants called by Lancet/Mutect2/Vardict, and indels within a 50bp window of a single gene. We excluded upstream and downstream gene variants from the analysis. We also excluded insertions and indels >5bp in length that were called by Vardict only because the caller does not output BAMs, preventing visual validation of the calls in Integrative Genomics Viewer (IGV). Smaller insertions and indels could be reviewed using BAM files from the alignment by BWA.

From the clean set of variants, we selected those that were annotated as pathogenic, likely-pathogenic, or pathogenic/likely pathogenic in ClinVar.^16^ We used AutoGVP^17^ to resolve variants with conflicting interpretations of pathogenicity. We also selected protein-coding variants annotated as oncogenic or likely oncogenic by OncoKB. Additionally, we developed custom criteria to profile variants without ClinVar or OncoKB records (Figure S2). We used ANNOVAR^18^ (November 2023 release) to annotate our data with gnomAD v.4.1.0^19^ allele frequencies and selected variants with allele frequencies ≤ 0.5%, excluding variants annotated benign or likely-benign in ClinVar, or neutral or likely neutral in OncoKB. Of these variants, we selected stop-gained, frameshift, splicing (SpliceAI delta score ≥ 0.5), and start-lost variants, accounting for escape of nonsense-mediated decay for those within tumor suppressor genes (TSG) (as assigned in COSMIC and OncoKB).^20,21^

#### Bioinformatics Analysis: Calculation of tumor mutational burden (TMB) and single-base substitution signatures (SBS)

Tumor mutational burden (TMB) was calculated according to the following formula: 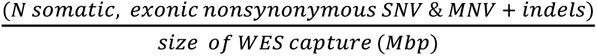 and single-base substitution (SBS) signatures (COSMIC) and their contributions to each tumor sample were identified using deconstructSigs.^22,23^ Tobacco smoking/chewing signatures, unknown signatures, and possible sequencing artefacts, as described by COSMIC, were not included in the contribution calculation.^22^ Only variants with alternative allele depth > 20, alternative allele frequency > 0.01, Varlociraptor^12^ somatic variant probability ≥ 0.9, and allele frequency ≤ 0.005 in gnomAD v.4.1.0 were used for determining TMB and SBS.

#### Bioinformatic Analysis: Identification of somatic pathogenic CNV, determination of allele-specific loss of heterozygosity, and calculation of homologous recombination deficiency

Allele-specific copy number variants (CNV) were called by a combination of ASCAT (v.3.1.3), FACETS (v.0.6.2), CNVkit (v.0.9.11)/PureCN (v.2.4.0) with segmentation by CNVkit and allele-specific calls by PureCN, and Sequenza (v.3.0), and they were annotated by AnnotSV (v.3.4.2).^24–29^ We filtered CNV by cancer genes and sorted by TSG and oncogenes as assigned in COSMIC and OncoKB. We called CNV variants in the TCGA samples using only FACETS and CNVkit.

In TSG, we selected biallelic loss, as defined by copy number 0 in both alleles, noted by at least 2 callers. In the oncogenes, we selected gain (total copy number = 3 or 4) and amplification (total copy number ≥ 5) with a consensus from at least 3 callers. When consensus was not reached, we calculated the average copy number and designated calls as 1) biallelic loss if average copy number was <0.5, 2) gain if average CN was >2.5 and <4.5, and 3) amplification if average total copy number was ≥4.5.

To evaluate the CNV landscape by chromosomal arm, we obtained genomic coordinates for each arm from the UCSC genome browser. The average total copy number from ASCAT, FACETS, PureCN, and Sequenza was calculated for each arm. We designated the calling for each arm based on the following rules: 1) loss if average total copy number was <1.5, 2) gain if average total CN was ≥2.5. The visualization is representative of the ASCAT segmental calls in IGV.

To evaluate joint loss of *RB1-BRCA2* and *TP53-BRCA1*, annotated allele-specific copy number variation (CNV) segments (including genomic positions and allele-specific copy numbers) generated by ASCAT were compiled for *BRCA1*, *BRCA2*, *RB1* and *TP53* in each tumor sample. For all tumors with allele-specific loss of heterozygosity in *RB1* or *TP53*, we noted the genomic coordinates of the beginning and end of the segment with copy loss that overlapped with each gene. We then determined how many times each lost segment overlapped with *RB1* only or *RB1* and *BRCA2*, or with *TP53* only, or *TP53* and *BRCA1*. We performed Chi-square tests to determine whether joint loss occurred more frequently than expected by chance.

Allele-specific LOH at *BRCA1/2* was noted if the tumor contained 1) a second somatic PV in the gene with the germline PV; or 2) allele-specific copy loss of the wildtype allele called by at least two out of four CNV callers used: ASCAT, FACETS, PureCN, and Sequenza. CNVs were annotated using AnnotSV.^24–28^

Genomic scars HRD score, defined as the sum of genomic LOH, telomeric allelic imbalance, and large state transitions, was calculated using HRDex with LOH values averaged from ASCAT, PureCN and Sequenza used as input.^30^

## Supplemental Tables (attached excel spreadsheet)

Table S1: Characteristics of *BRCA1/2* pathogenic variant carriers in the full POSH study and in the current study

Table S2: List of cancer genes evaluated

Table S3: Homologous recombination deficiency scores and tumor mutational burden in tumors

Table S4: Single base substitution signatures in tumors

Table S5: Frequency of variants identified in tumors

Table S6: Significantly altered genes by gene variant

Table S7: Significantly altered genes by estrogen receptor status

Table S8: Significantly altered genes in homologous recombination deficiency high and low tumors

Table S9: Significantly altered genes in tumors with and without allele-specific loss of heterozygosity

## Supplemental Figures

**Figure S1:**
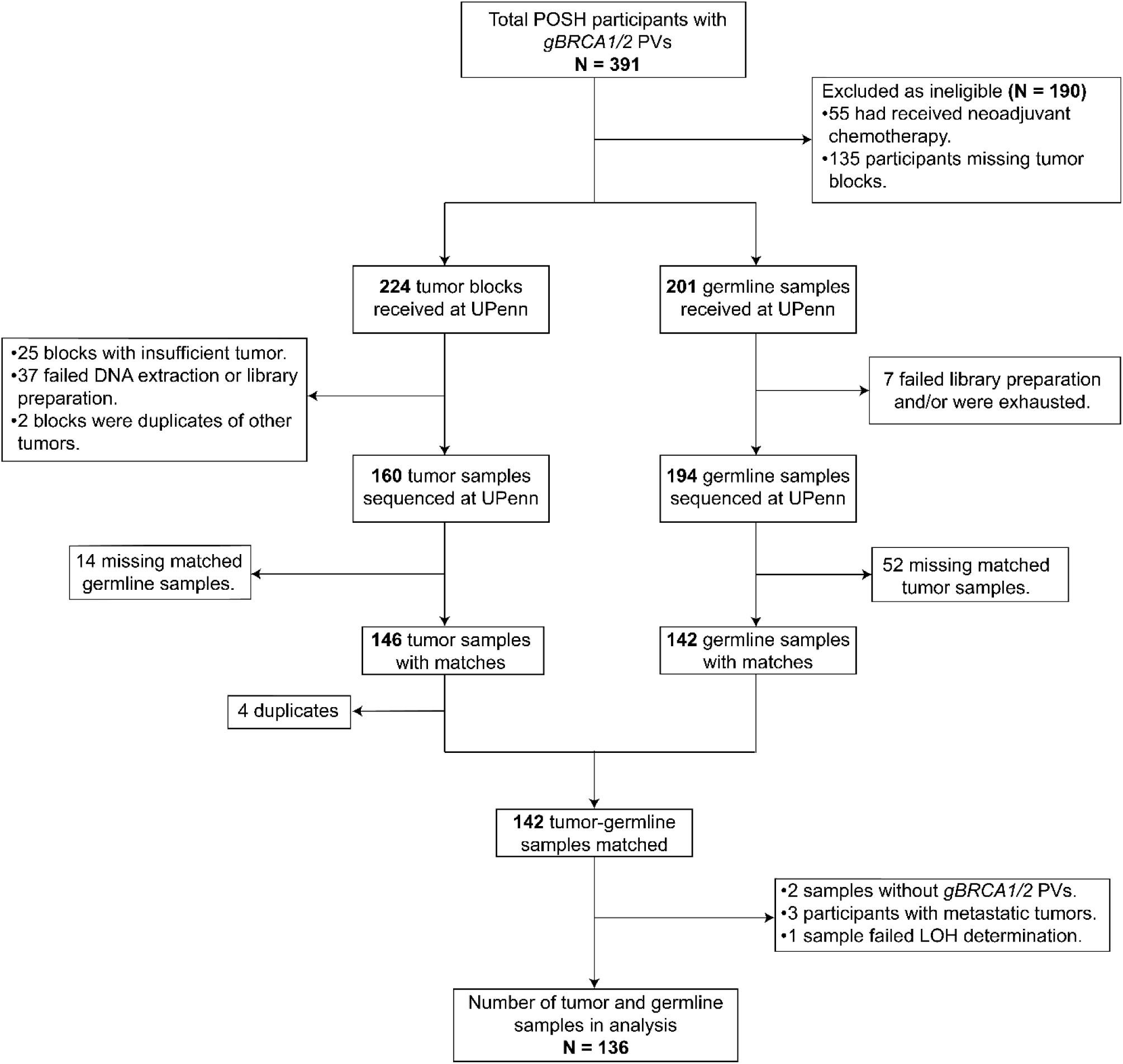
CONSORT diagram of POSH study samples

**Figure S2:**
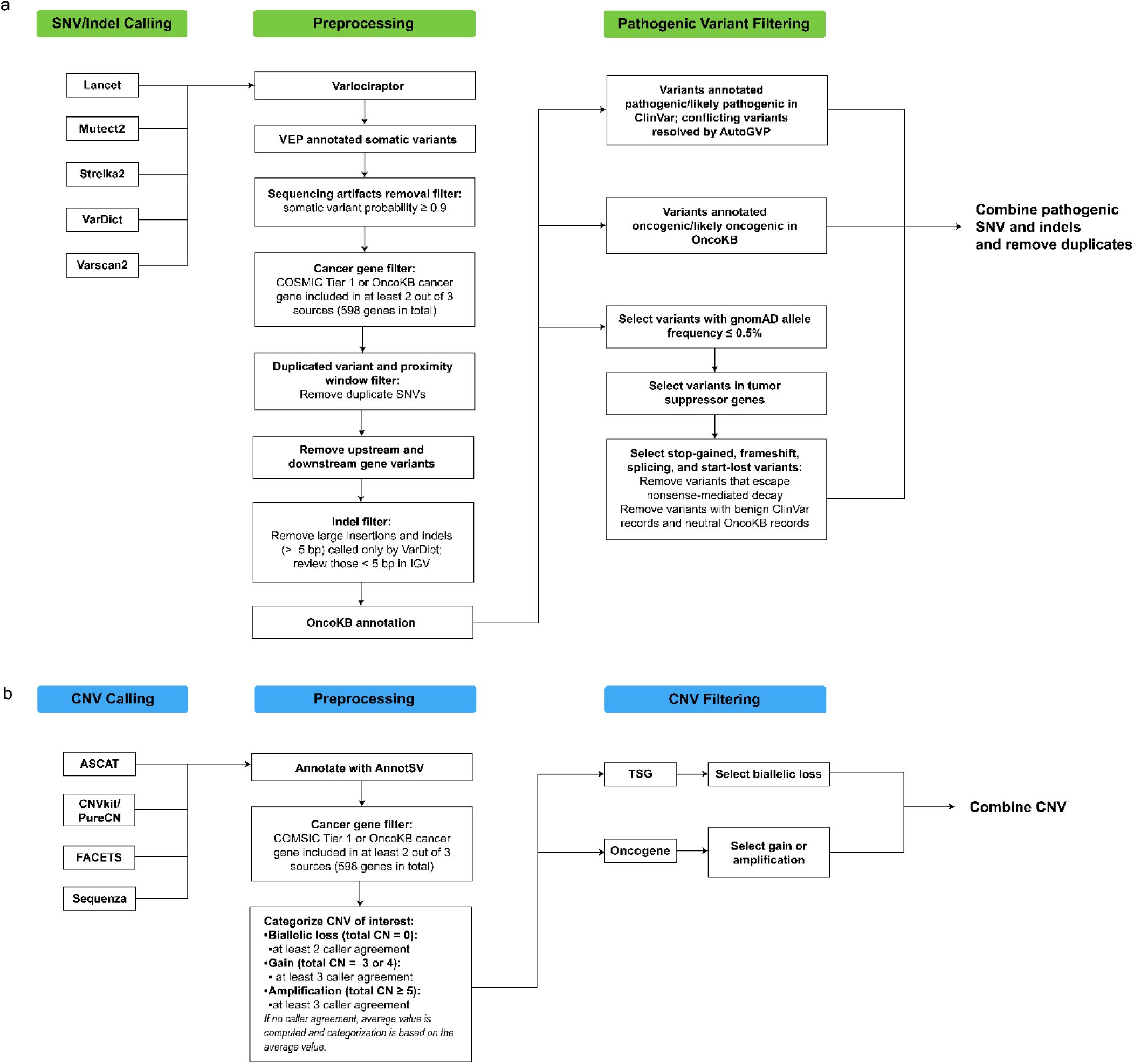
Custom somatic variant analysis pipeline.

**Figure S3:**
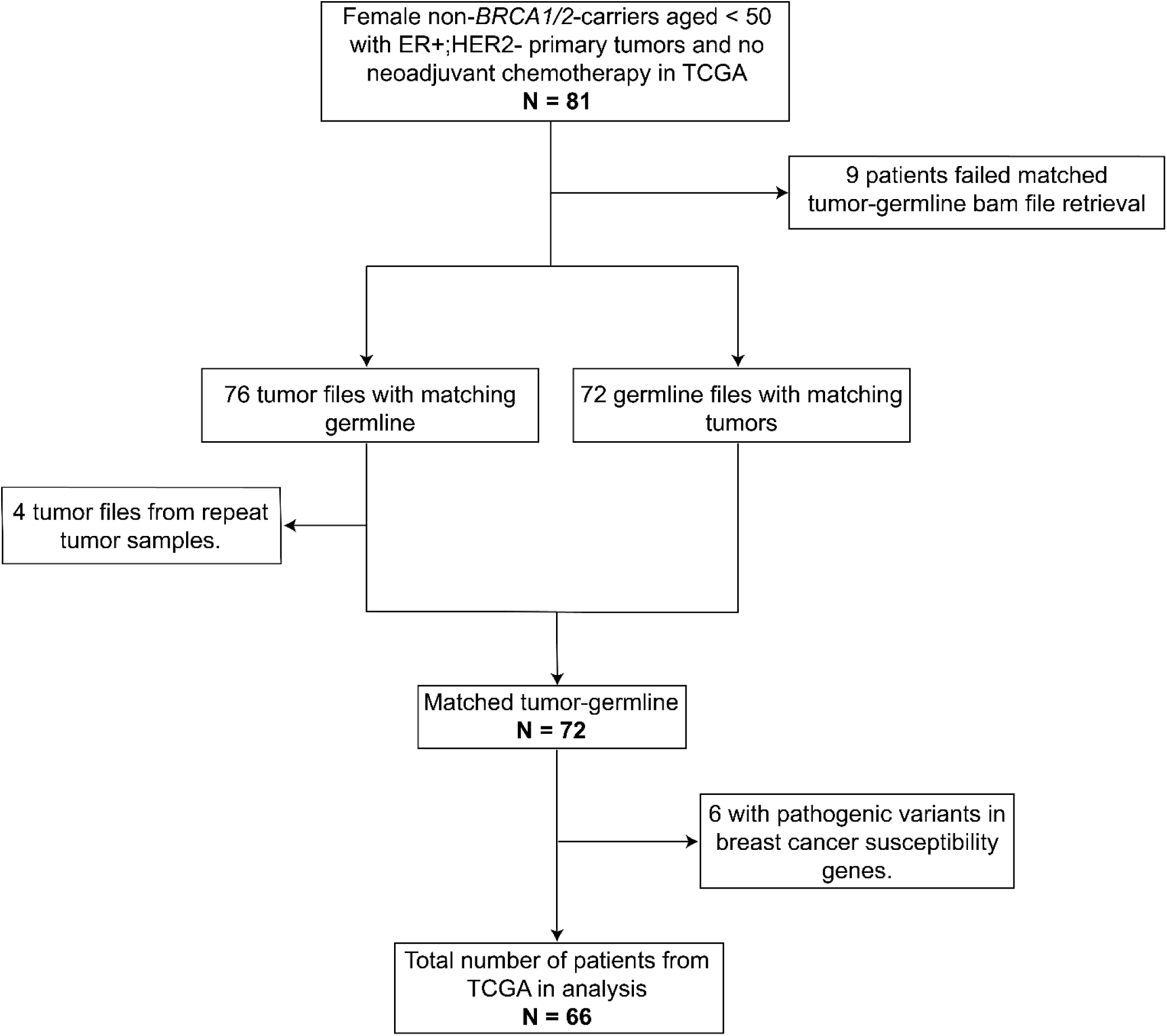
CONSORT diagram of TCGA samples used in this study.

**Figure S4:**
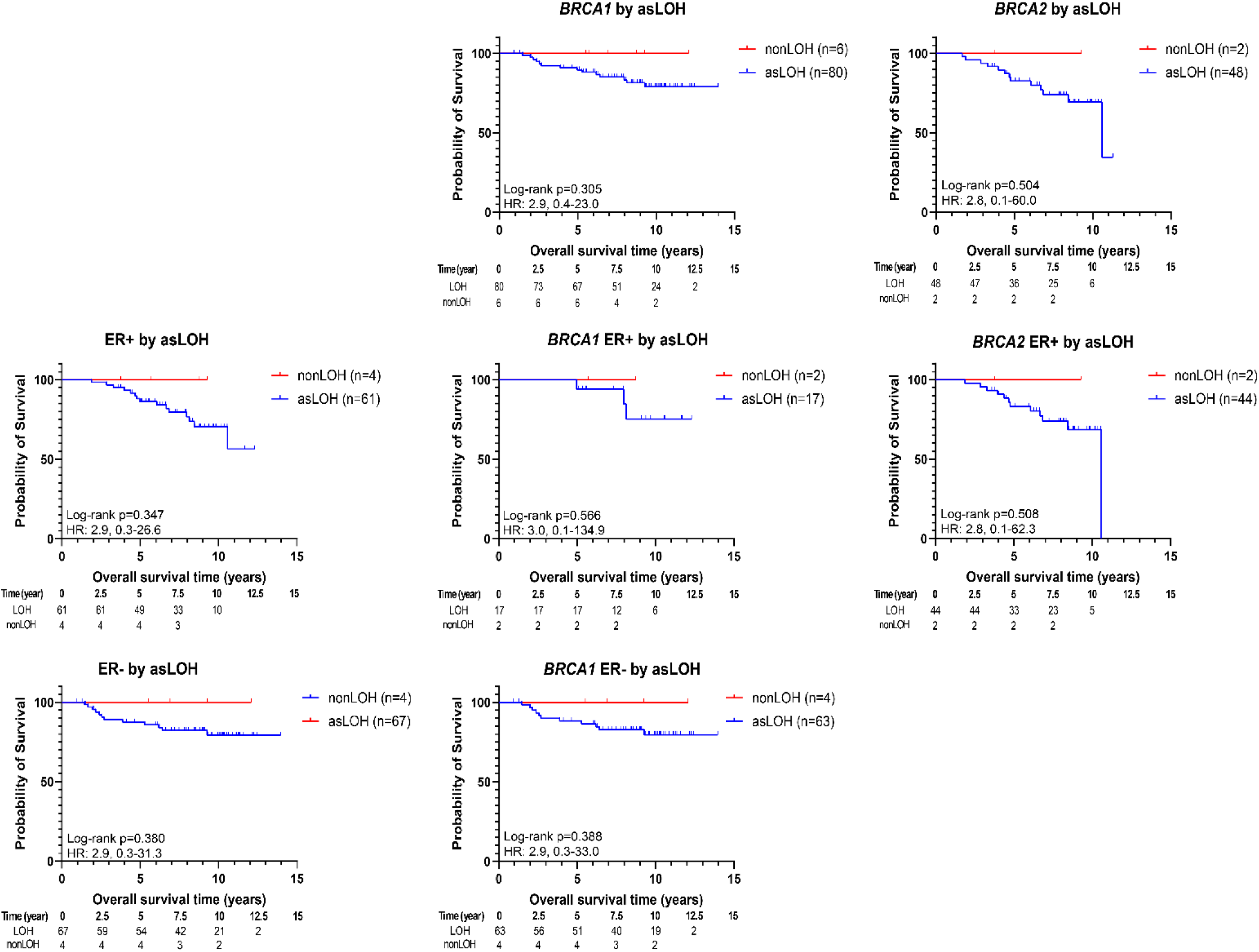
Overall survival by loss of heterozygosity status.

**Figure S5:**
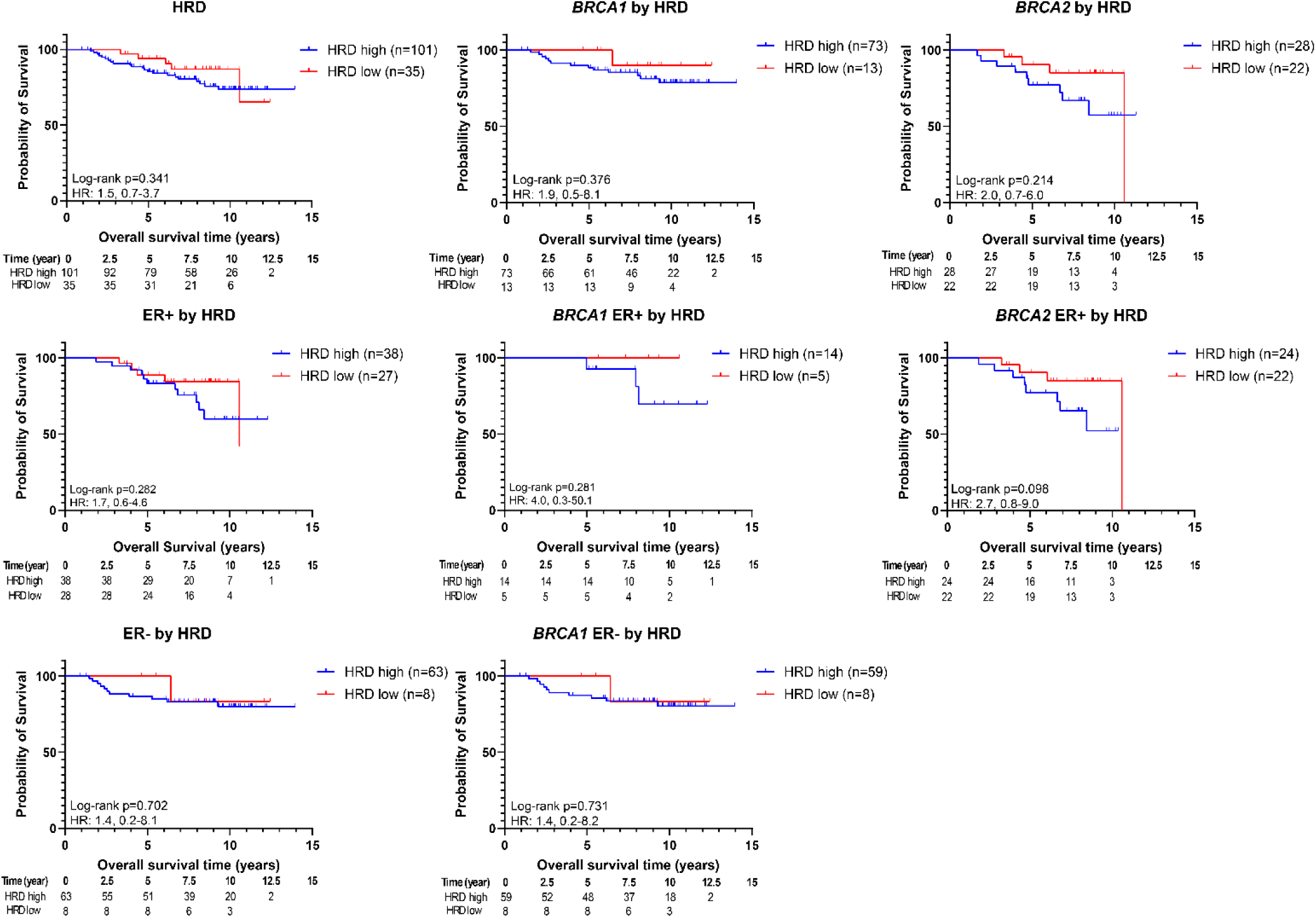
Overall survival by homologous recombination deficiency (HRD).

**Figure S6:**
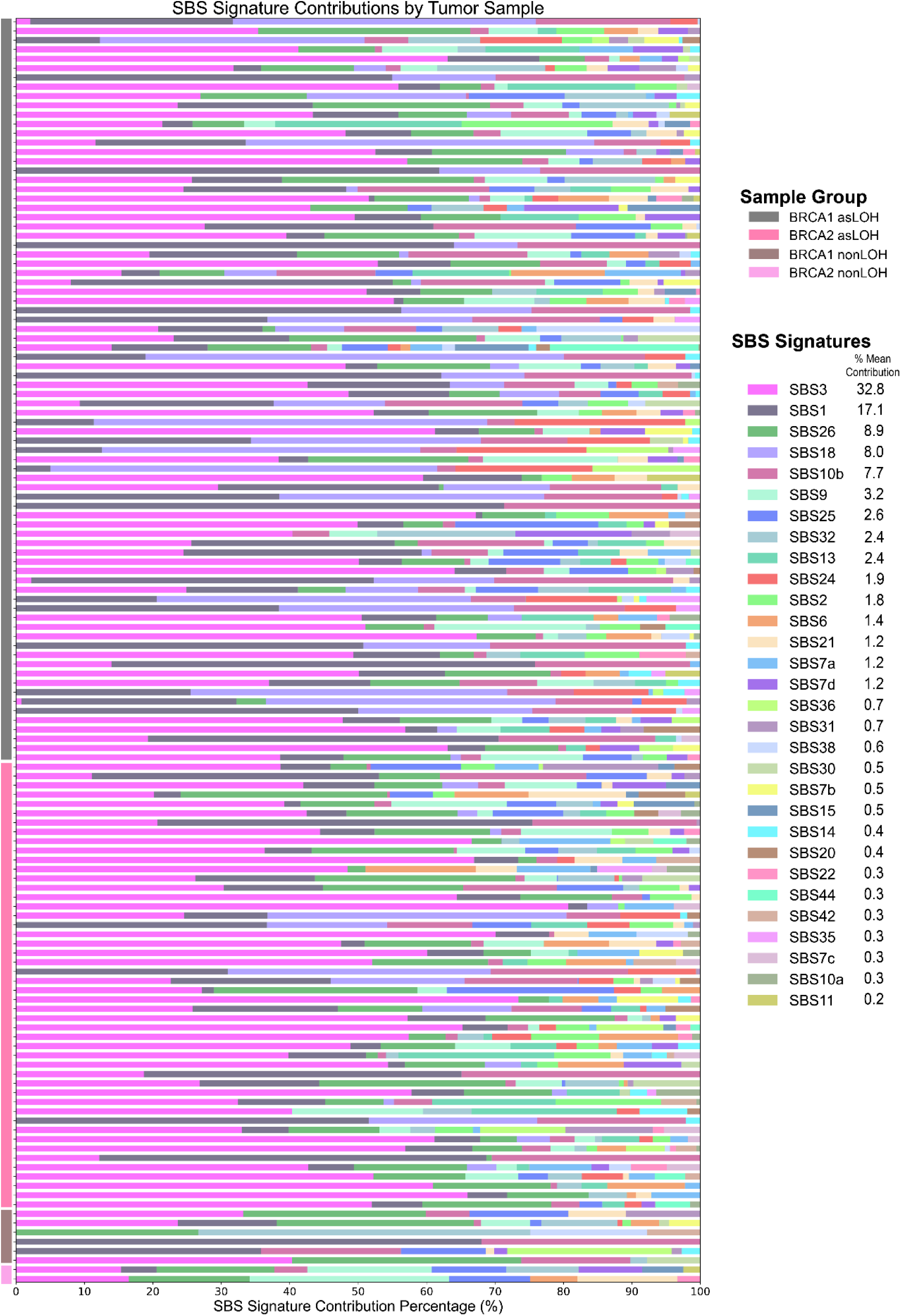
Landscape of single base substitutions in tumors in POSH.

**Figure S7:**
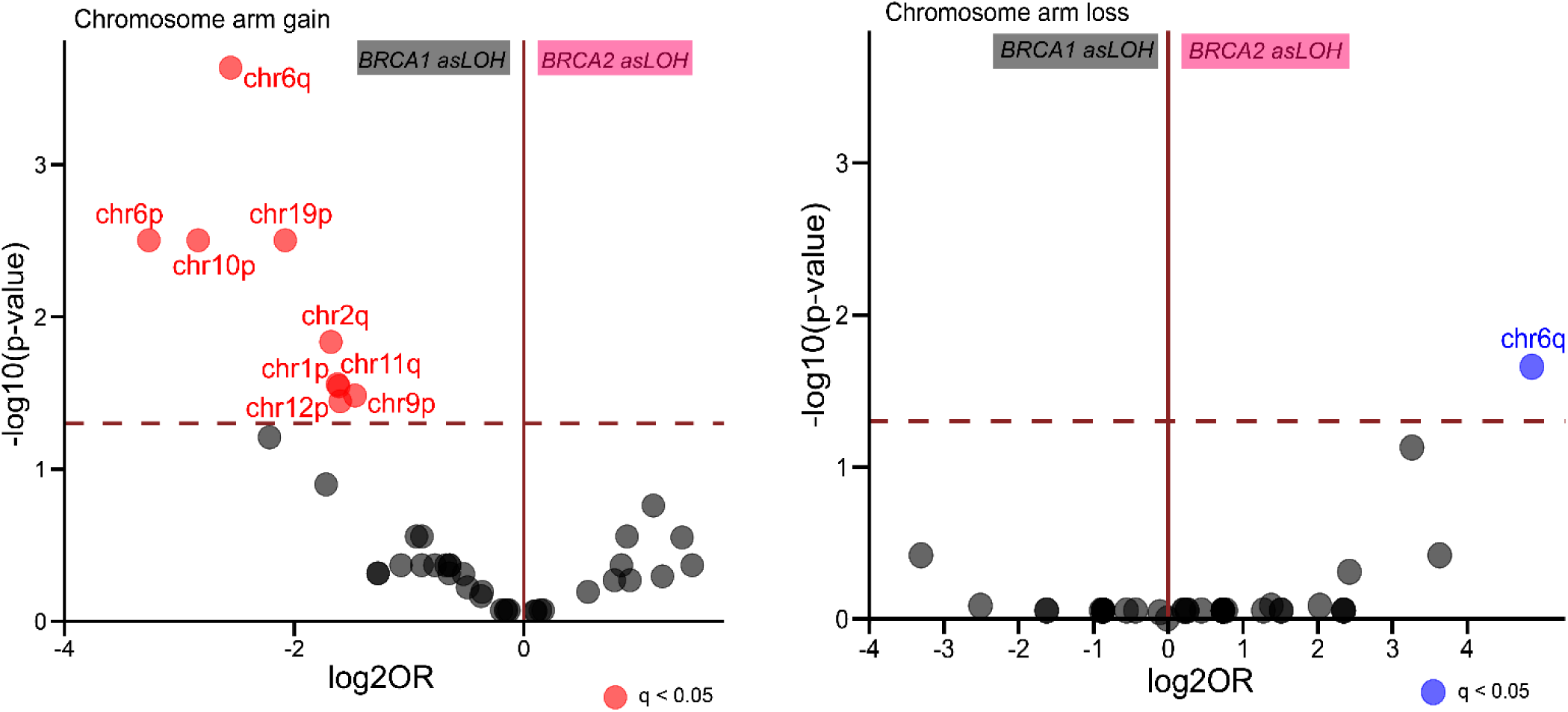
Differential enrichment of chromosome arm copy changes in *BRCA1* and *BRCA2* tumors with asLOH adjusted for multiple testing.

**Figure S8:**
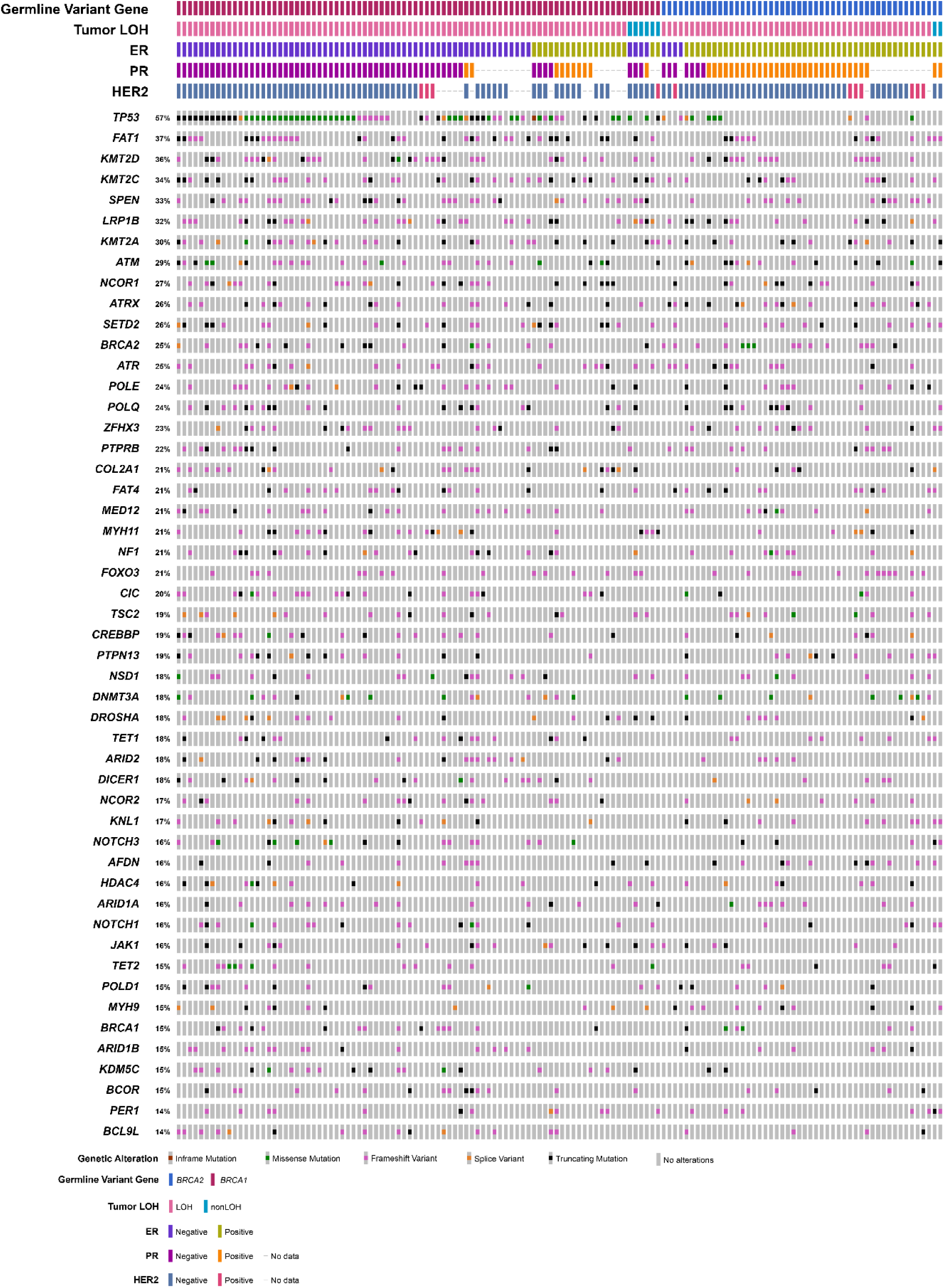
Oncoprint of the 50 most altered genes by single nucleotide variants and indels.

**Figure S9:**
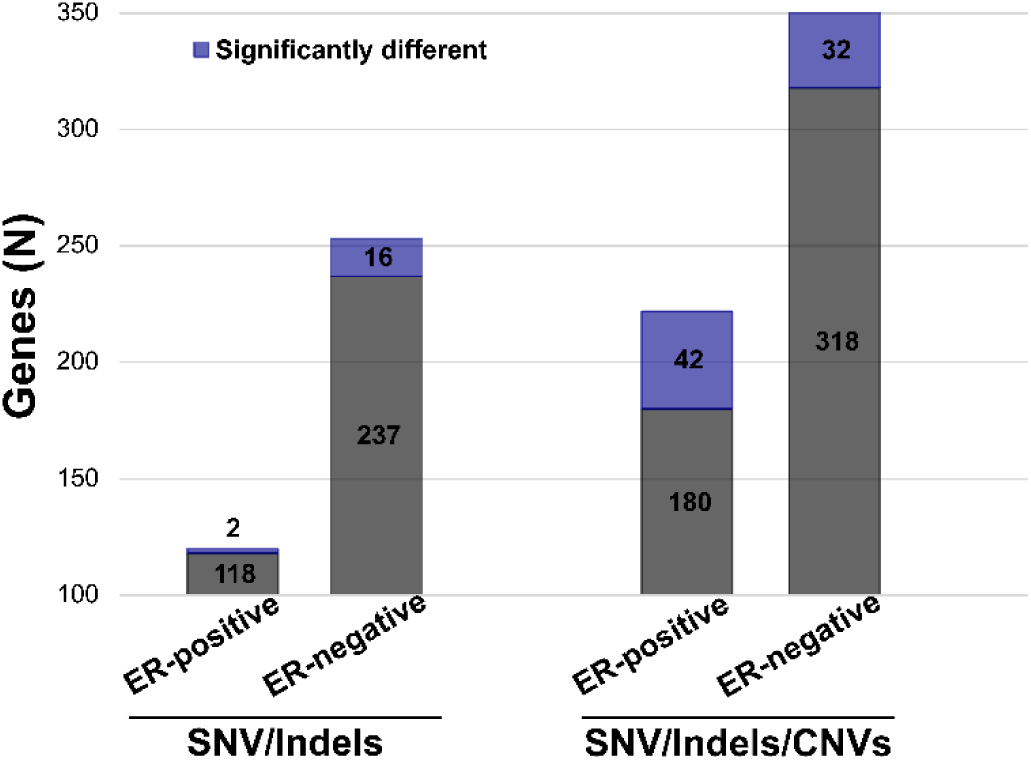
Number of genes with single nucleotide variants/indels/copy number variants by estrogen-receptor status.

## References

1. Armstrong N, Ryder S, Forbes C, et al: A systematic review of the international prevalence of BRCA mutation in breast cancer. Clin Epidemiol 11:543–561, 2019

2. Kurian AW, Ward KC, Howlader N, et al: Genetic Testing and Results in a Population-Based Cohort of Breast Cancer Patients and Ovarian Cancer Patients. J Clin Oncol 37:1305–1315, 2019

3. Copson ER, Maishman TC, Tapper WJ, et al: Germline BRCA mutation and outcome in young-onset breast cancer (POSH): a prospective cohort study. Lancet Oncol 19:169–180, 2018

4. Evans DG, van Veen EM, Byers HJ, et al: High likelihood of actionable pathogenic variant detection in breast cancer genes in women with very early onset breast cancer. Journal of Medical Genetics 59:115–121, 2022

5. Dorling L, Carvalho S, Allen J, et al: Breast Cancer Risk Genes — Association Analysis in More than 113,000 Women. New England Journal of Medicine 384:428–439, 2021

6. Mavaddat N, Peock S, Frost D, et al: Cancer Risks for BRCA1 and BRCA2 Mutation Carriers: Results From Prospective Analysis of EMBRACE. JNCI: Journal of the National Cancer Institute 105:812–822, 2013

7. Shubeck S, Sevilimedu V, Berger E, et al: Comparison of Outcomes Between BRCA Pathogenic Variant Carriers Undergoing Breast-Conserving Surgery Versus Mastectomy. Ann Surg Oncol 29:4706–4713, 2022

8. Valachis A, Nearchou AD, Lind P: Surgical management of breast cancer in BRCA-mutation carriers: a systematic review and meta-analysis. Breast Cancer Res Treat 144:443–55, 2014

9. Hu C, Hart SN, Gnanaolivu R, et al: A Population-Based Study of Genes Previously Implicated in Breast Cancer. New England Journal of Medicine 384:440–451, 2021

10. Moynahan ME, Chiu JW, Koller BH, et al: Brca1 controls homology-directed DNA repair. Mol Cell 4:511–8, 1999

11. Moynahan ME, Pierce AJ, Jasin M: BRCA2 is required for homology-directed repair of chromosomal breaks. Mol Cell 7:263–72, 2001

12. Maxwell KN, Wubbenhorst B, Wenz BM, et al: BRCA locus-specific loss of heterozygosity in germline BRCA1 and BRCA2 carriers. Nature Communications 8:319, 2017

13. Mavaddat N, Barrowdale D, Andrulis IL, et al: Pathology of Breast and Ovarian Cancers among BRCA1 and BRCA2 Mutation Carriers: Results from the Consortium of Investigators of Modifiers of BRCA1/2 (CIMBA). Cancer Epidemiology, Biomarkers & Prevention 21:134–147, 2012

14. Consortium BCA: Pathology of Tumors Associated With Pathogenic Germline Variants in 9 Breast Cancer Susceptibility Genes. JAMA Oncology 8:e216744–e216744, 2022

15. Inagaki-Kawata Y, Yoshida K, Kawaguchi-Sakita N, et al: Genetic and clinical landscape of breast cancers with germline BRCA1/2 variants. Communications Biology 2020 3:1 3, 2020-10-16

16. Palacios J, Robles Frías MJ, Castilla MA, et al: The Molecular Pathology of Hereditary Breast Cancer. Pathobiology 75, 2008/06/01

17. Lal A, Ramazzotti D, Weng Z, et al: Comprehensive genomic characterization of breast tumors with BRCA1 and BRCA2 mutations. BMC Medical Genomics 2019 12:1 12, 2019-06-10

18. Samstein RM, Krishna C, Ma X, et al: Mutations in BRCA1 and BRCA2 differentially affect the tumor microenvironment and response to checkpoint blockade immunotherapy. Nature Cancer 2020 1:12 1, 2020-11-16

19. van Beers EH, van Welsem T, Wessels LFA, et al: Comparative Genomic Hybridization Profiles in Human BRCA1 and BRCA2 Breast Tumors Highlight Differential Sets of Genomic Aberrations. Cancer Research 65, 2005/02/01

20. Melchor L, Honrado E, Huang J, et al: Estrogen Receptor Status Could Modulate the Genomic Pattern in Familial and Sporadic Breast Cancer. Clinical Cancer Research 13:7305–7313, 2007

21. Tirkkonen M, Johannsson O, Agnarsson BA, et al: Distinct somatic genetic changes associated with tumor progression in carriers of BRCA1 and BRCA2 germ-line mutations. Cancer Res 57:1222–7, 1997

22. Hedenfalk I, Duggan D, Chen Y, et al: Gene-Expression Profiles in Hereditary Breast Cancer. New England Journal of Medicine 344:539–548, 2001

23. Farmer H, McCabe N, Lord CJ, et al: Targeting the DNA repair defect in BRCA mutant cells as a therapeutic strategy. Nature 434:917–21, 2005

24. Go RS, Adjei AA: Review of the comparative pharmacology and clinical activity of cisplatin and carboplatin. J Clin Oncol 17:409–22, 1999

25. Litton JK, Rugo HS, Ettl J, et al: Talazoparib in Patients with Advanced Breast Cancer and a Germline *BRCA* Mutation. New England Journal of Medicine 379:753–763, 2018

26. Robson M, Im SA, Senkus E, et al: Olaparib for Metastatic Breast Cancer in Patients with a Germline BRCA Mutation. N Engl J Med 377:523–533, 2017

27. Tutt ANJ, Garber JE, Kaufman B, et al: Adjuvant Olaparib for Patients with BRCA1- or BRCA2-Mutated Breast Cancer. N Engl J Med 384:2394–2405, 2021

28. Geyer CE, Jr., Garber JE, Gelber RD, et al: Overall survival in the OlympiA phase III trial of adjuvant olaparib in patients with germline pathogenic variants in BRCA1/2 and high-risk, early breast cancer. Ann Oncol 33:1250–1268, 2022

29. Johnston SRD, Harbeck N, Hegg R, et al: Abemaciclib Combined With Endocrine Therapy for the Adjuvant Treatment of HR+, HER2-, Node-Positive, High-Risk, Early Breast Cancer (monarchE). J Clin Oncol 38:3987–3998, 2020

30. Johnston SRD, Toi M, O’Shaughnessy J, et al: Abemaciclib plus endocrine therapy for hormone receptor-positive, HER2-negative, node-positive, high-risk early breast cancer (monarchE): results from a preplanned interim analysis of a randomised, open-label, phase 3 trial. Lancet Oncol 24:77–90, 2023

31. Slamon D, Lipatov O, Nowecki Z, et al: Ribociclib plus Endocrine Therapy in Early Breast Cancer. N Engl J Med 390:1080–1091, 2024

32. Im S-A, Lu Y-S, Bardia A, et al: Overall Survival with Ribociclib plus Endocrine Therapy in Breast Cancer. New England Journal of Medicine 381:307–316, 2019

33. Kaufman B, Shapira-Frommer R, Schmutzler RK, et al: Olaparib monotherapy in patients with advanced cancer and a germline BRCA1/2 mutation. J Clin Oncol 33:244–50, 2015

34. Lord CJ, Ashworth A: Mechanisms of resistance to therapies targeting BRCA-mutant cancers. Nat Med 19:1381–8, 2013

35. Nik-Zainal S, Davies H, Staaf J, et al: Landscape of somatic mutations in 560 breast cancer whole-genome sequences. Nature 534:47–54, 2016

36. Tutt A, Tovey H, Cheang MCU, et al: Carboplatin in BRCA1/2-mutated and triple-negative breast cancer BRCAness subgroups: the TNT Trial. Nature Medicine 24:628–637, 2018

37. Li Q, Jiang B, Guo J, et al: INK4 Tumor Suppressor Proteins Mediate Resistance to CDK4/6 Kinase Inhibitors. Cancer Discovery 12:356–371, 2022/02/01

38. Wander SA, Cohen O, Gong X, et al: The Genomic Landscape of Intrinsic and Acquired Resistance to Cyclin-Dependent Kinase 4/6 Inhibitors in Patients with Hormone Receptor– Positive Metastatic Breast Cancer. Cancer Discovery 10:1174–1193, 2020/08/01

39. Ganesan S: Tumor Suppressor Tolerance: Reversion Mutations in BRCA1 and BRCA2 and Resistance to PARP Inhibitors and Platinum. JCO Precision Oncology:1–4, 2018

40. Wineland D, Le AN, Hausler R, et al: Biallelic BRCA Loss and Homologous Recombination Deficiency in Nonbreast/Ovarian Tumors in Germline BRCA1/2 Carriers. JCO Precis Oncol 7:e2300036, 2023

41. Ding L, Cao J, Lin W, et al: The Roles of Cyclin-Dependent Kinases in Cell-Cycle Progression and Therapeutic Strategies in Human Breast Cancer. Int J Mol Sci 21, 2020

42. Wang X, Zhao S, Xin Q, et al: Recent progress of CDK4/6 inhibitors’ current practice in breast cancer. Cancer Gene Ther 31:1283–1291, 2024

43. Goetz MP, Hamilton EP, Campone M, et al: Landscape of Baseline and Acquired Genomic Alterations in Circulating Tumor DNA with Abemaciclib Alone or with Endocrine Therapy in Advanced Breast Cancer. Clinical Cancer Research 30, 2024/05/15

44. Wander SA, Cohen O, Gong X, et al: The Genomic Landscape of Intrinsic and Acquired Resistance to Cyclin-Dependent Kinase 4/6 Inhibitors in Patients with Hormone Receptor– Positive Metastatic Breast Cancer. Cancer Discovery 10, 2020/08/01

45. Eccles D, Gerty S, Simmonds P, et al: Prospective study of Outcomes in Sporadic versus Hereditary breast cancer (POSH): study protocol. BMC Cancer 7:160, 2007

46. Schouten JP, McElgunn CJ, Waaijer R, et al: Relative quantification of 40 nucleic acid sequences by multiplex ligation-dependent probe amplification. Nucleic Acids Res 30:e57, 2002

47. Heath AP, Ferretti V, Agrawal S, et al: The NCI Genomic Data Commons. Nature Genetics 53:257–262, 2021

48. Telli ML, Timms KM, Reid J, et al: Homologous Recombination Deficiency (HRD) Score Predicts Response to Platinum-Containing Neoadjuvant Chemotherapy in Patients with Triple-Negative Breast Cancer. Clinical Cancer Research 22:3764–3773, 2016

49. Liberzon A, Birger C, Thorvaldsdóttir H, et al: The Molecular Signatures Database (MSigDB) hallmark gene set collection. Cell Syst 1:417–425, 2015

50. Shahrouzi P, Forouz F, Mathelier A, et al: Copy number alterations: a catastrophic orchestration of the breast cancer genome. Trends in Molecular Medicine 30:750–764, 2024

51. Li Z, Zhang X, Hou C, et al: Comprehensive identification and characterization of somatic copy number alterations in triple-negative breast cancer. Int J Oncol 56:522–530, 2020

52. Pereira B, Chin S-F, Rueda OM, et al: The somatic mutation profiles of 2,433 breast cancers refine their genomic and transcriptomic landscapes. Nature Communications 7:11479, 2016

53. Goetz MP, Hamilton EP, Campone M, et al: Landscape of Baseline and Acquired Genomic Alterations in Circulating Tumor DNA with Abemaciclib Alone or with Endocrine Therapy in Advanced Breast Cancer. Clinical Cancer Research 30:2233–2244, 2024/05/15

54. Li Z, Razavi P, Li Q, et al: Loss of the FAT1 Tumor Suppressor Promotes Resistance to CDK4/6 Inhibitors via the Hippo Pathway. Cancer Cell 34:893–905.e8, 2018

55. Morrison L, Loibl S, Turner NC: The CDK4/6 inhibitor revolution — a game-changing era for breast cancer treatment. Nature Reviews Clinical Oncology 21:89–105, 2024

56. Safonov A, Marra A, Bandlamudi C, et al: Homologous recombination deficiency and tumor suppressor heterozygosity mediate resistance to front-line therapy in breast cancer. bioRxiv:2024.02.05.578934, 2024

57. André F, Solovieff N, Su F, et al: Acquired gene alterations in patients treated with ribociclib plus endocrine therapy or endocrine therapy alone using baseline and end-of-treatment circulating tumor DNA samples in the MONALEESA-2, -3, and -7 trials. Annals of Oncology 36:54–64, 2024

58. van Geelen CT, Teo ZL, Savas P, et al: Abstract P5-02-16: SPEN is a biomarker for CDK4/6 inhibitor resistance in patients with metastatic hormone receptor positive (HR+)/HER2-breast cancer. Cancer Research 83:P5-02-16-P5-02-16, 2023

59. Alexandrov LB, Kim J, Haradhvala NJ, et al: The repertoire of mutational signatures in human cancer. Nature 578:94–101, 2020

60. Jönsson Gr, Naylor TL, Vallon-Christersson J, et al: Distinct Genomic Profiles in Hereditary Breast Tumors Identified by Array-Based Comparative Genomic Hybridization. Cancer Research 65:7612–7621, 2005

61. Kauraniemi Pi, Hedenfalk I, Persson K, et al: MYB Oncogene Amplification in Hereditary BRCA1 Breast Cancer1. Cancer Research 60:5323–5328, 2000

62. O’Neil SR, Weber GA, Deming DA, et al: Exceptional Response to Crizotinib With Subsequent Response to Cabozantinib in Metastatic, ROS1-GOPC Fusion–Mutated Breast Cancer. JCO Precision Oncology:e2300174, 2023

63. Hornsveld M, Smits LMM, Meerlo M, et al: FOXO Transcription Factors Both Suppress and Support Breast Cancer Progression. Cancer Research 78:2356–2369, 2018

64. Lu M, Hartmann D, Braren R, et al: Oncogenic Akt-FOXO3 loop favors tumor-promoting modes and enhances oxidative damage-associated hepatocellular carcinogenesis. BMC Cancer 19:887, 2019

65. Ter Steege EJ, Boer M, Timmer NC, et al: R-spondin-3 is an oncogenic driver of poorly differentiated invasive breast cancer. J Pathol 258:289–299, 2022

66. Bai F, Wang C, Liu X, et al: Loss of function of BRCA1 promotes EMT in mammary tumors through activation of TGFβR2 signaling pathway. Cell Death & Disease 13:195, 2022

67. Gorrini C, Baniasadi PS, Harris IS, et al: BRCA1 interacts with Nrf2 to regulate antioxidant signaling and cell survival. Journal of Experimental Medicine 210:1529–1544, 2013

68. Saha T, Rih JK, Rosen EM: BRCA1 down-regulates cellular levels of reactive oxygen species. FEBS Lett 583:1535–43, 2009

69. Bajrami I, Marlow R, van de Ven M, et al: E-Cadherin/ROS1 Inhibitor Synthetic Lethality in Breast Cancer. Cancer Discov 8:498–515, 2018

70. Fischer MM, Yeung VP, Cattaruzza F, et al: RSPO3 antagonism inhibits growth and tumorigenicity in colorectal tumors harboring common Wnt pathway mutations. Sci Rep 7:15270, 2017

71. Bendell J, Eckhardt GS, Hochster HS, et al: 68 - Initial results from a phase 1a/b study of OMP-131R10, a first-in-class anti-RSPO3 antibody, in advanced solid tumors and previously treated metastatic colorectal cancer (CRC). European Journal of Cancer 69:S29–S30, 2016

72. Inagaki-Kawata Y, Yoshida K, Kawaguchi-Sakita N, et al: Genetic and clinical landscape of breast cancers with germline BRCA1/2 variants. Communications Biology 3:578, 2020

73. Rouault A, Banneau G, MacGrogan G, et al: Deletion of Chromosomes 13q and 14q Is a Common Feature of Tumors with BRCA2 Mutations. PLOS ONE 7:e52079, 2012

74. Martins FC, De S, Almendro V, et al: Evolutionary Pathways in BRCA1-Associated Breast Tumors. Cancer Discovery 2:503–511, 2012

75. Bruno L, Ostinelli A, Waisberg F, et al: Cyclin-Dependent Kinase 4/6 Inhibitor Outcomes in Patients With Advanced Breast Cancer Carrying Germline Pathogenic Variants in DNA Repair–Related Genes. JCO Precision Oncology 6:e2100140, 2022

76. Kim J-Y, Oh JM, Park YH, et al: Which Clinicopathologic Parameters Suggest Primary Resistance to Palbociclib in Combination With Letrozole as the First-Line Treatment for Hormone Receptor-Positive, HER2-Negative Advanced Breast Cancer? Frontiers in Oncology 11:759150, 2021

## Supplemental references

1. Li H, Durbin R: Fast and accurate short read alignment with Burrows–Wheeler transform. Bioinformatics 25:1754–60, 2009/07/15

2. Danecek P, Bonfield JK, Liddle J, et al: Twelve years of SAMtools and BCFtools. GigaScience 10, 2021/01/29

3. Poplin R, Chang P-C, Alexander D, et al: A universal SNP and small-indel variant caller using deep neural networks. Nature Biotechnology 2018 36:10 36:983-987, 2018-09-24

4. Lai Z, Markovets A, Ahdesmaki M, et al: VarDict: a novel and versatile variant caller for next-generation sequencing in cancer research. Nucleic Acids Research 44:e108, 2016/06/20

5. Anderson CA, Pettersson FH, Clarke GM, et al: Data quality control in genetic case-control association studies. Nature Protocols 5:1564–1573, 2010

6. Purcell S, Neale B, Todd-Brown K, et al: PLINK: A Tool Set for Whole-Genome Association and Population-Based Linkage Analyses. The American Journal of Human Genetics 81:559–75, 2007/09/01

7. Narzisi G, Corvelo A, Arora K, et al: Genome-wide somatic variant calling using localized colored de Bruijn graphs. Communications Biology 2018 1:1 1:20, 2018-03-22

8. Benjamin D, Sato T, Cibulskis K, et al: Calling Somatic SNVs and Indels with Mutect2. bioRxiv, 2019-12-02

9. Kim S, Scheffler K, Halpern AL, et al: Strelka2: fast and accurate calling of germline and somatic variants. Nature Methods 2018 15:8 15:591-594, 2018-07-16

10. Koboldt DC, Zhang Q, Larson DE, et al: VarScan 2: Somatic mutation and copy number alteration discovery in cancer by exome sequencing. Genome Research 22:568–76, 2012-03-01

11. McLaren W, Gil L, Hunt SE, et al: The Ensembl Variant Effect Predictor. Genome Biology 2016 17:1 17:122, 2016-06-06

12. Köster J, Dijkstra LJ, Marschall T, et al: Varlociraptor: enhancing sensitivity and controlling false discovery rate in somatic indel discovery. Genome Biology 2020 21:1 21:98, 2020-04-28

13. Sondka Z, Dhir NB, Carvalho-Silva D, et al: COSMIC: a curated database of somatic variants and clinical data for cancer. Nucleic Acids Research 52:D1210–D1217, 2024/01/05

14. Chakravarty D, Gao J, Phillips S, et al: OncoKB: A Precision Oncology Knowledge Base. JCO Precision Oncology 1, 2017-05-16

15. Suehnholz SP, Nissan MH, Zhang H, et al: Quantifying the Expanding Landscape of Clinical Actionability for Patients with Cancer. Cancer Discovery 14:49–65, 2024/01/01

16. Landrum MJ, Lee JM, Riley GR, et al: ClinVar: public archive of relationships among sequence variation and human phenotype. Nucleic Acids Research 42:D980–5, 2014/01/01

17. Kim J, Naqvi AS, Corbett RJ, et al: AutoGVP: a dockerized workflow integrating ClinVar and InterVar germline sequence variant classification. Bioinformatics 40:btae114, 2024

18. Wang K, Li M, Hakonarson H: ANNOVAR: functional annotation of genetic variants from high-throughput sequencing data. Nucleic Acids Research 38:e164, 2010/09/01

19. Karczewski KJ, Francioli LC, Tiao G, et al: The mutational constraint spectrum quantified from variation in 141,456 humans. Nature 2020 581:7809 581:434-443, 2020-05-27

20. Jaganathan K, Kyriazopoulou Panagiotopoulou S, McRae JF, et al: Predicting Splicing from Primary Sequence with Deep Learning. Cell 176:535–548.e24, 2019

21. Lindeboom RGH, Supek F, Lehner B, et al: The rules and impact of nonsense-mediated mRNA decay in human cancers. Nature Genetics 2016 48:10 48:1112-8, 2016-09-12

22. Alexandrov LB, Kim J, Haradhvala NJ, et al: The repertoire of mutational signatures in human cancer. Nature 578:94–101, 2020

23. Rosenthal R, McGranahan N, Herrero J, et al: deconstructSigs: delineating mutational processes in single tumors distinguishes DNA repair deficiencies and patterns of carcinoma evolution. Genome Biology 2016 17:1 17:31, 2016-02-22

24. Ross EM, Haase K, Van Loo P, et al: Allele-specific multi-sample copy number segmentation in ASCAT. Bioinformatics 37:1909–1911, 2021/07/27

25. Shen R, Seshan VE: FACETS: allele-specific copy number and clonal heterogeneity analysis tool for high-throughput DNA sequencing. Nucleic Acids Research 44:e131, 2016/09/19

26. Riester M, Singh AP, Brannon AR, et al: PureCN: copy number calling and SNV classification using targeted short read sequencing. Source Code for Biology and Medicine 2016 11:1 11:13, 2016-12-15

27. Favero F, Joshi T, Marquard AM, et al: Sequenza: allele-specific copy number and mutation profiles from tumor sequencing data. Annals of Oncology 26:64–70, 2015/01/01

28. Geoffroy V, Herenger Y, Kress A, et al: AnnotSV: an integrated tool for structural variations annotation. Bioinformatics 34:3572–3574, 2018/10/15

29. Talevich E, Shain AH, Botton T, et al: CNVkit: Genome-Wide Copy Number Detection and Visualization from Targeted DNA Sequencing. PLoS Comput Biol 12:e1004873, 2016

30. Pluta J, Hausler R, Wubbenhorst B, et al: HRDex: a tool for deriving homologous recombination deficiency (HRD) scores from whole exome sequencing data. bioRxiv, 2022-09-12

